# Empowering patients across the maternal-newborn care continuum: A cluster randomized controlled trial testing a digital health platform in Kenya

**DOI:** 10.1101/2024.06.03.24308340

**Authors:** Rajet Vatsa, Wei Chang, Sharon Akinyi, Sarah Little, Catherine Gakii, John Mungai, Cynthia Kahumbura, Anneka Wickramanayake, Sathy Rajasekharan, Jessica Cohen, Margaret McConnell

## Abstract

**Background:** Accelerating improvements in maternal and newborn healthcare is a major public health priority in Kenya. While utilization of formal healthcare has increased, many pregnant and postpartum women do not receive the recommended number of maternal care visits. Even when they do, visits are often short with many providers not offering important elements of evaluation and counseling, leaving gaps in women’s knowledge and preparedness. Digital health tools have been proposed as a complement to care that is provided by maternity care facilities, with the potential to empower patients to receive the right care at the right place and time. However, there is limited evidence of the impact of digital health tools at scale on patients’ knowledge, preparedness, and the content of care they receive. We evaluated a digital health platform (PROMPTS) composed of informational messages, appointment reminders, and a two-way clinical helpdesk that has been implemented at scale in Kenya on six domains across the pregnancy-postpartum care continuum.

**Methods and Findings:** We conducted an unmasked, 1:1 parallel arm cluster randomized controlled trial in 40 health facilities (clusters) across eight counties in Kenya. 6,139 pregnant individuals were consented at baseline and followed through pregnancy and postpartum.

Individuals recruited from treatment facilities were invited to enroll in the PROMPTS platform, with roughly 85% reporting take-up. Our outcomes were derived from phone surveys conducted with participants at 36-42 weeks of gestation and 7-8 weeks post-childbirth. Among eligible participants, 3,399/3,678 women completed antenatal follow-up, and 5,509/6,128 women completed postpartum follow-up, with response rates of 92% and 90%, respectively. Outcomes were organized into six domains: knowledge, preparedness, routine care seeking, danger sign care seeking, newborn care, and postpartum care content. We generated standardized summary indices to account for multiple hypothesis testing but also analyzed individual index components.

Intention-to-treat analyses were conducted for all outcomes at the individual level, with standard errors clustered by facility. Participants recruited from treatment facilities had a 0.08 standard deviation (SD) (95% CI: 0.03, 0.12) higher knowledge index, a 0.08 SD (95% CI: 0.02, 0.13) higher preparedness index, a 0.10 SD (95% CI: 0.05, 0.16) higher routine care seeking index, a 0.09 SD (95% CI: 0.07, 0.12) higher newborn care index, and a 0.06 SD (95% CI: 0.01, 0.12) higher postpartum care content index than those recruited from control facilities. No significant effect on the danger sign care seeking index was found (95% CI: -0.01, 0.08).

A limitation of our study was that outcomes were self-reported, and the study was not powered to detect effects on health outcomes.

**Conclusions:** Digital health tools indicate promise in filling gaps in pregnant and postpartum women’s health care, amidst systems that fail to deliver a minimally adequate standard of care. Through providing patients with critical information and empowering them to seek recommended care, such tools can ensure that individuals are prepared for a safe childbirth and receive access to comprehensive, high quality postpartum care. Future work is needed to ascertain the impact of at-scale digital platforms like PROMPTS on health outcomes.

**Trial Registration:** ClinicalTrials.gov ID: NCT05110521; AEA RCT Registry ID: [R-0008449](https://www.socialscienceregistry.org/trials/8449)

## INTRODUCTION

While maternal and newborn health (MNH) outcomes have improved globally, progress over the past decade has been slow or stagnant in many low- and lower-middle-income settings. In Kenya, for example, the maternal mortality rate remains well over five times the United Nations’ Sustainable Development Goal.^1^ Stillbirth and neonatal mortality rates in Kenya have also been stagnant for much of the 21^st^ century, though declines in neonatal mortality have been reported in recent years.^2,3^ In addition, high rates of maternal and neonatal severe morbidity persist across sub-Saharan Africa, including in Kenya.^4,5,6,7^

Accelerating improvements in MNH care to reduce mortality and morbidity is a major public health priority in Kenya. In recent decades, there have been substantial shifts in utilization of formal maternity care: 90% of pregnant women in Kenya receive some form of prenatal care from a medical professional and over 80% of births occur at health facilities, nearly double the rate from 2003.^8,9,10^ Nevertheless, 40% of pregnant women do not receive the recommended number of antenatal care (ANC) visits, and just over 50% receive any postpartum care, rates that have remained stable over the past two decades.10^,11,12^ In some regions of Kenya, it has also been noted that the content of postpartum care focuses on the needs of the newborn exclusively, rather than on those of both the mother and newborn.^13^ Moreover, when visits do happen, studies from low- and-middle-income settings have revealed that time spent with providers is often short, frequently lasting less than five minutes.^14^ A recent multi-country survey with respondents from Kenya relatedly found that ∼1/3 of patients reported inadequate time with their providers.^15^

Amidst this backdrop of mixed advances towards adequate formal care utilization, research increasingly highlights the need for improvements in the quality of care.^15,16,17^ However, existing evidence from Kenya suggests that the majority of women do not receive a minimally adequate standard of antenatal and postpartum care.^18^ For example, many patients are not offered a host of essential care components, with systematic patient assessments revealing that – across poverty levels – fewer than 60% of individuals were comprehensively examined or counseled on critical topics like pregnancy complications and birth preparation.^15,18^ Thus, many pregnant and postpartum women remain ill-informed about key aspects of their health and are not empowered to seek recommended care – for both routine and acute cases – at the right place and time.10^,19,20,21^ When patients are underequipped with such knowledge, it can result in delays in the decision to seek care. These delayed care decisions are often referred to as the “first delay” in the Three Delays Model of MNH care and are associated with increased maternal and neonatal morbidity and mortality.^19,22,23,24^

Historically, such challenges have been addressed at both the health system level and patient level. Health system level strategies predominantly entail some form of provider training; however, these approaches can be costly and have demonstrated mixed success, with modest effect sizes and issues with sustainability at scale.^15,25,26,27,28^ Patient level strategies, on the other hand, include programs that mitigate barriers to care seeking (e.g., support groups, transport vouchers, insurance) and informational tools that promote health literacy and receipt of recommended care.^27,28^ Such approaches have demonstrated success in improving care utilization but can be challenging to integrate with the formal MNH care system. Moreover, to date, they have underemphasized outcomes for mothers in the postpartum period.^27,29,30^

Digital health tools, however, offer unique promise as a patient level strategy to complement care provided in the formal sector and close the gaps described previously across the pregnancy-postpartum continuum. With high rates of mobile phone penetration across low-income countries, the prevalence of such interventions has increased in recent decades.^31^ To date, digital health tools targeting MNH care have primarily sought to promote recommended health behaviors and care seeking through SMS- or voice-based education and nudges.^30,32,33^ Other tools aim to connect patients with care providers via digital communication platforms (e.g., digital helpdesks).32 In multiple systematic reviews, existing tools have been associated with improvements in utilization of antenatal care, facility-based birth, skilled birth attendance, and newborn vaccination, though few studies have examined or identified an impact on knowledge, preparedness, or content of care, particularly for mothers in the postpartum setting.^30,31,32^

With the recent rise of artificial intelligence (AI) as well, such tools can be layered with AI to efficiently triage problems and deliver targeted education to patients.^34^ Thus, if integrated effectively, digital health tools have the potential to reduce strain on the formal healthcare system, enhance access to critical information, and empower patients to receive the right care and the right place and time. Ultimately, well-powered, randomized evidence from low- and middle-income settings on the impact of digital health tools on a broad set of MNH outcomes is critical to understand how these programs could scale in implementation settings.^31,32,35^

In this paper, we describe a parallel arm cluster randomized controlled trial (RCT) carried out in 40 health facilities across Kenya to test the impact of a digital health platform for targeted patient communication. The platform, PROMPTS (Promoting Mothers in Pregnancy and Postpartum Through SMS), was developed by Jacaranda Health, a leading MNH nonprofit in Kenya, to “empower women to seek care at the right time and place and give them greater agency in the health system.”^36^ Through collaborations with regional and national government officials, over 1,200 health facilities across Kenya have begun enrolling pregnant and postpartum women onto the platform, with over two million individuals having enrolled as of November 2023. Accordingly, our study contributes to the literature by rigorously evaluating the impact of a patient level solution for MNH care that has been implemented at scale. In addition, we expand beyond the narrow focus on guideline-recommended antenatal and postnatal care seeking and examine the effect of PROMPTS on knowledge, preparedness, and content of care, for both pregnant and postpartum women and their newborns.

## METHODS

We conducted an unmasked, 1:1 parallel arm cluster RCT in 40 health facilities across eight counties in Kenya (**Fig 1**). Treatment facilities received two different interventions, one “patient-facing” and the other “provider-facing”. The first was that women attending antenatal care at treatment facilities were invited to enroll in PROMPTS. The second was a nurse mentorship program designed to increase and sustain providers’ knowledge and skills in basic and emergency obstetric and newborn care during the childbirth hospitalization. We evaluated these components separately, focusing here on the impact of facilities’ offering PROMPTS on pregnant and postpartum women’s knowledge, preparedness, care utilization, health behaviors, and content of care, using longitudinal surveys of participants across the prenatal and postpartum settings. The nurse mentorship program was focused largely on quality of emergency care during childbirth and was not intended to influence the outcomes analyzed here. Further, 97% of the PROMPTS sample delivered in study facilities before rollout of the nurse training was complete. Finally, none of the outcomes analyzed in the PROMPTS evaluation could be influenced by the other intervention. The nurse training intervention is being evaluated in separate work.

**Fig 1:**
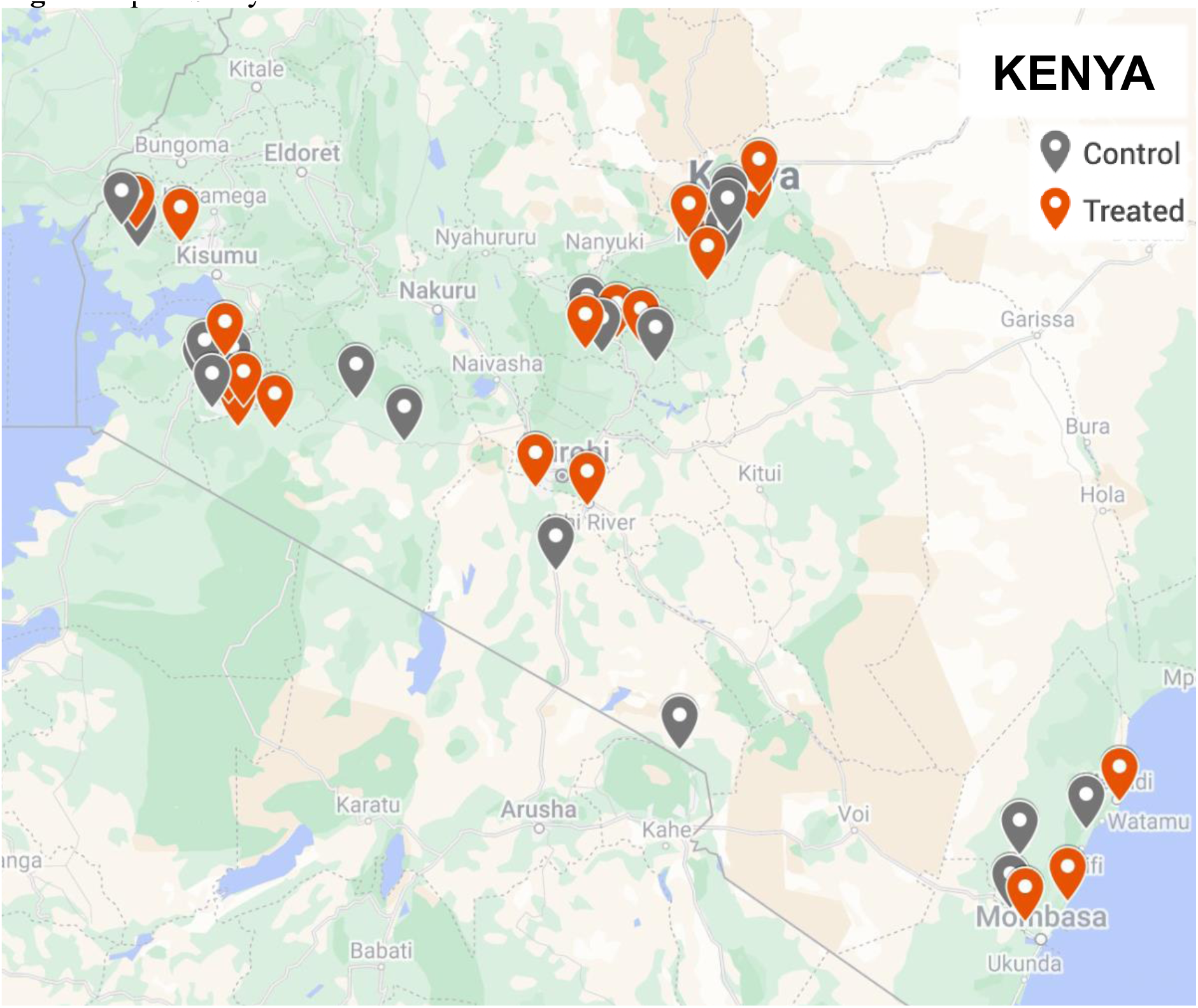
Map of Study Facilities. This figure displays the location of the 40 health facilities in the study. The study facilities are distributed across eight counties (Kajiado [four facilities], Kilifi [six], Kirinyaga [two], Kisii [eight], Meru [eight], Narok [four], Nyeri [four], and Siaya [four]). Facilities are color-coded by treatment vs. control status. For 33 of the 40 study facilities, the closest facility is 10+ kilometers away. For the remaining seven facilities, the nearest facility belongs to the same study arm. The average minimum distance between facilities of opposite arms is 32 kilometers.

Patients receiving prenatal care at treatment facilities were invited to enroll in PROMPTS during in-person interactions with onsite enumerators. In control facilities, PROMPTS was not systematically introduced to participants, either during interactions with study enumerators or through standard processes of care. However, women enrolled from control facilities were unrestricted from independently enrolling in PROMPTS and may have done so through other channels, such as learning about the platform from personal contacts or providers exposed to it. Evaluation outcomes were measured through longitudinal surveys with women during the prenatal and postpartum period.

We pre-registered our evaluation of the complete intervention package on the United States Clinical Trials Registry (NCT05110521) and the American Economic Association’s RCT Registry (AEARCTR-0008449).

### Intervention

PROMPTS consists of informational messages, appointment reminders, and a two-way clinical helpdesk. SMS messages in English or Swahili – determined based on language preferences solicited at enrollment – are sent to enrollees based on their gestational age and are designed to influence relevant health behaviors. The content that participants receive has been rigorously tested with intended beneficiaries by Jacaranda Health and accordingly adapted to ensure broad accessibility. Depending on the timing of enrollment, participants are sent roughly 10-40 messages per month during pregnancy and roughly 10 messages per month during the first year postpartum. The PROMPTS platform also includes an AI-enabled helpdesk that assesses, triages, and responds to questions. The helpdesk first runs messages through a natural language processing pipeline to assign each message an intent and priority level. Accordingly, the helpdesk sends automated guidance (roughly 86% of cases) or escalates messages to trained clinical agents who staff the helpdesk 24/7 and respond within one minute for high-priority cases.^36,37^ Finally, PROMPTS users are queried about their maternal care content and experience and the aggregated data is shared with facilities and health system managers to build accountability and target system improvements. PROMPTS is free for participants, runs via SMS on both basic and smartphones, and is primarily rolled out through public health facilities.

### Procedures

#### Health Facility Eligibility

Health facilities were eligible for study inclusion if they were owned by the government or a faith-based organization and had an average of 50-400 normal (i.e., unassisted) vaginal deliveries per month, based on records from the Kenya Health Management Information System. Facilities were also eligible based on having no known ongoing mobile health or quality-improvement research initiatives. 37 of the 40 included study facilities were owned by the Ministry of Health of Kenya, and three were owned by faith-based organizations. The facilities were primarily Level 4 hospitals (32 facilities), though five were Level 3 health centers and three were Level 5 county referral hospitals. Facilities assigned to different study arms had a minimum distance of 10 kilometers between them to avoid spillovers, with an average minimum distance of 32 kilometers between facilities of opposite arms (**Fig 1**).

#### Randomization

Randomization was conducted at the facility level given that enrollment into PROMPTS is designed to take place at health facilities. Facility-level randomization was stratified by tertile of monthly volume of normal vaginal deliveries (see **Appendix S1** for details).

#### Recruitment and Eligibility

Pregnant women were recruited by onsite enumerators at health facilities during antenatal care and were followed up by phone during pregnancy and postpartum. To be eligible, participants had to have access to a mobile phone (basic phone or smartphone), be at least 15 years old, and be at least 16 weeks pregnant or in month 5-9 of pregnancy.

#### Data Collection

All participants completed an in-person informed consent process followed by a baseline survey. A subsample of women (those who were enrolled prior to their 36^th^ week of gestation) were contacted by mobile phone for a follow-up antenatal survey in the final weeks of pregnancy (i.e., around 36-42 weeks of gestation). All women surveyed at baseline were contacted by mobile phone for a follow-up postpartum survey, 7-8 weeks after childbirth. Participants received an SMS credit of 100 Kenyan shillings per survey completed.

Baseline and follow-up surveys asked questions about demographic and health information, healthcare seeking, knowledge of maternal and newborn danger signs, health practices and preparedness, and content of care. In cases of stillbirth, miscarriage, or infant loss at antenatal or postpartum follow-up, participants were offered resources to connect with trained psychological counselors and to abstain from being interviewed. In cases where participants elected to continue being interviewed, a shortened survey was administered. Baseline data collection lasted from November to December 2021, antenatal follow-up data collection lasted from November 2021 to May 2022, and postpartum follow-up data collection lasted from January to August 2022 (**Fig 2**).

**Fig 2:**
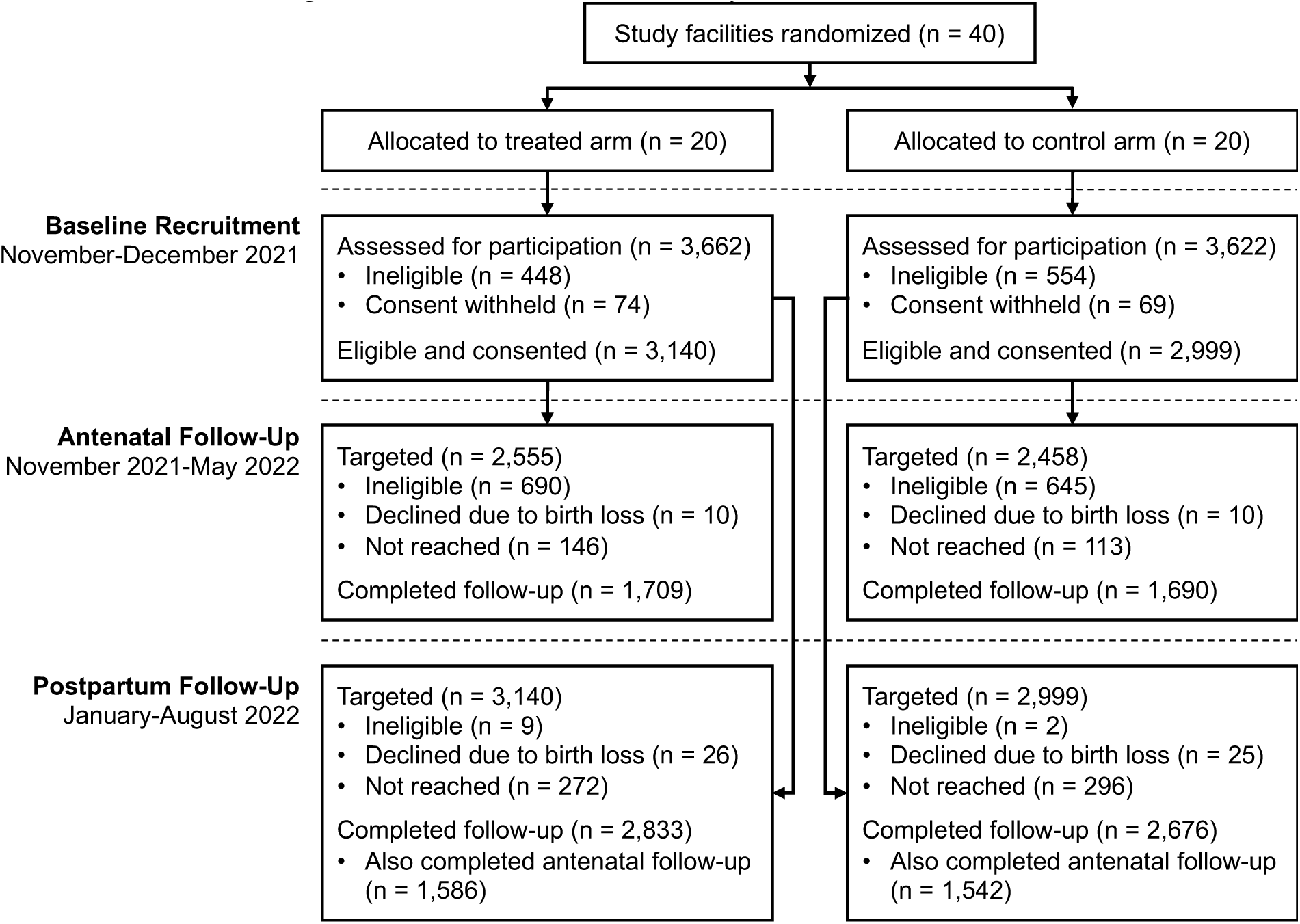
Retention at Antenatal and Postpartum Follow-Up of the Baseline Cohort of Pregnant Women Recruited During Antenatal Care Visits to Study Facilities. This figure synthesizes the baseline recruitment of pregnant women who visited health facilities for antenatal care and tracks their retention at antenatal and postpartum follow-up, which occurred in the final weeks of pregnancy (i.e., around 36-42 weeks of gestation) and 7-8 weeks post-childbirth, respectively. Women recruited at or beyond 36 weeks of gestation at baseline were not targeted for antenatal follow-up, while participants whose pregnancy had ended with birth of a living newborn were targeted but ineligible. Meanwhile, for postpartum follow-up, all eligible and consented participants at baseline were targeted and only a handful were ineligible due to being fewer than seven weeks postpartum.

#### Enrollment in PROMPTS

Immediately following the baseline survey, women in treatment facilities were invited to enroll in PROMPTS by study enumerators. At the time of consent and enrollment, women were provided with high-level information about the platform, an overview of the program’s goals to mitigate delays in care seeking, and an assurance of the study team’s commitment to data protection and privacy. Those who agreed to participate started receiving messages as soon as their contact information was shared with Jacaranda Health, typically within 24 hours of enrollment.

### Outcomes

We organized individual-level pre-registered primary and secondary outcomes into six domains: knowledge, preparedness, routine care seeking, danger sign care seeking, newborn care, and postpartum care content. **Fig 3** offers a conceptual framework motivating the assessment of outcomes in these domains. For domains with more than two outcomes, we generated summary indices to account for multiple hypothesis testing, à la Anderson (2008).^38^ Specifically, for each index, we ensured directional consistency of the component outcomes and calculated inverse-covariance-weighted averages of normalized versions of each component. In a few instances, we included exploratory outcomes that were not pre-registered but were strongly connected to the program’s theory of change; pre-registered vs. exploratory outcomes are clearly denoted throughout. In addition, **Table 1** presents a detailed set of PROMPTS messages that correspond to each outcome.

**Fig 3:**
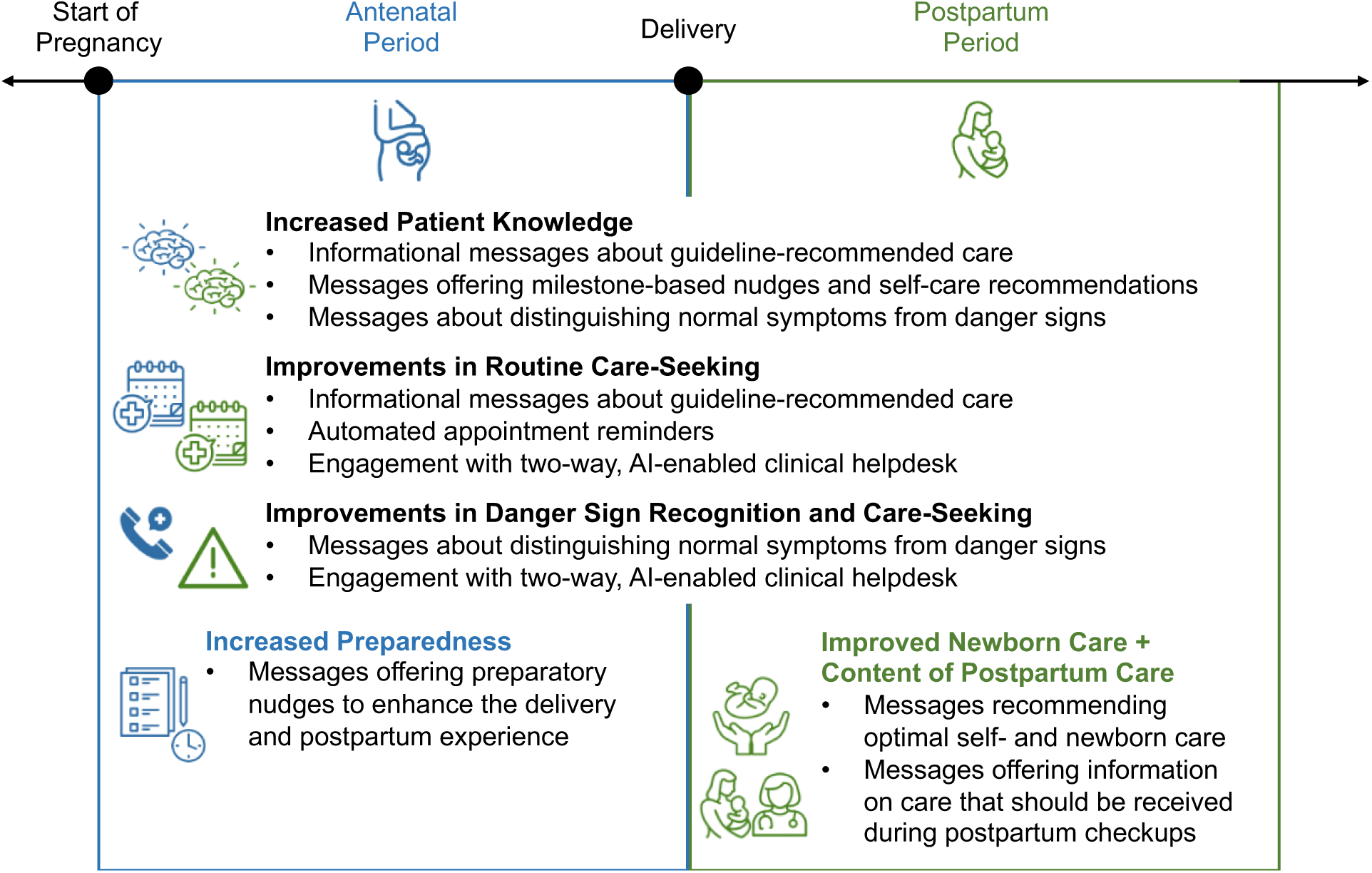
Conceptual Framework of the Domains Through Which PROMPTS May Have an Effect Across the Pregnancy-Postpartum Care Continuum. This figure offers a conceptual framework of the potential impacts PROMPTS could have. PROMPTS offers support across the pregnancy-postpartum care continuum. During both the antenatal and postpartum periods, improvements in patient knowledge (e.g., through informational messages) could lead to improvements in routine care seeking and danger sign recognition/care seeking. During the antenatal period, messages containing preparatory nudges may enhance preparation for childbirth, while during the postpartum period, messages containing care recommendations and content of care reminders may enhance newborn care and postpartum care content for infants and mothers. PROMPTS is also comprised of automated appointment reminders and a two-way clinical helpdesk, which may influence routine and danger sign care seeking.

**Table 1:**
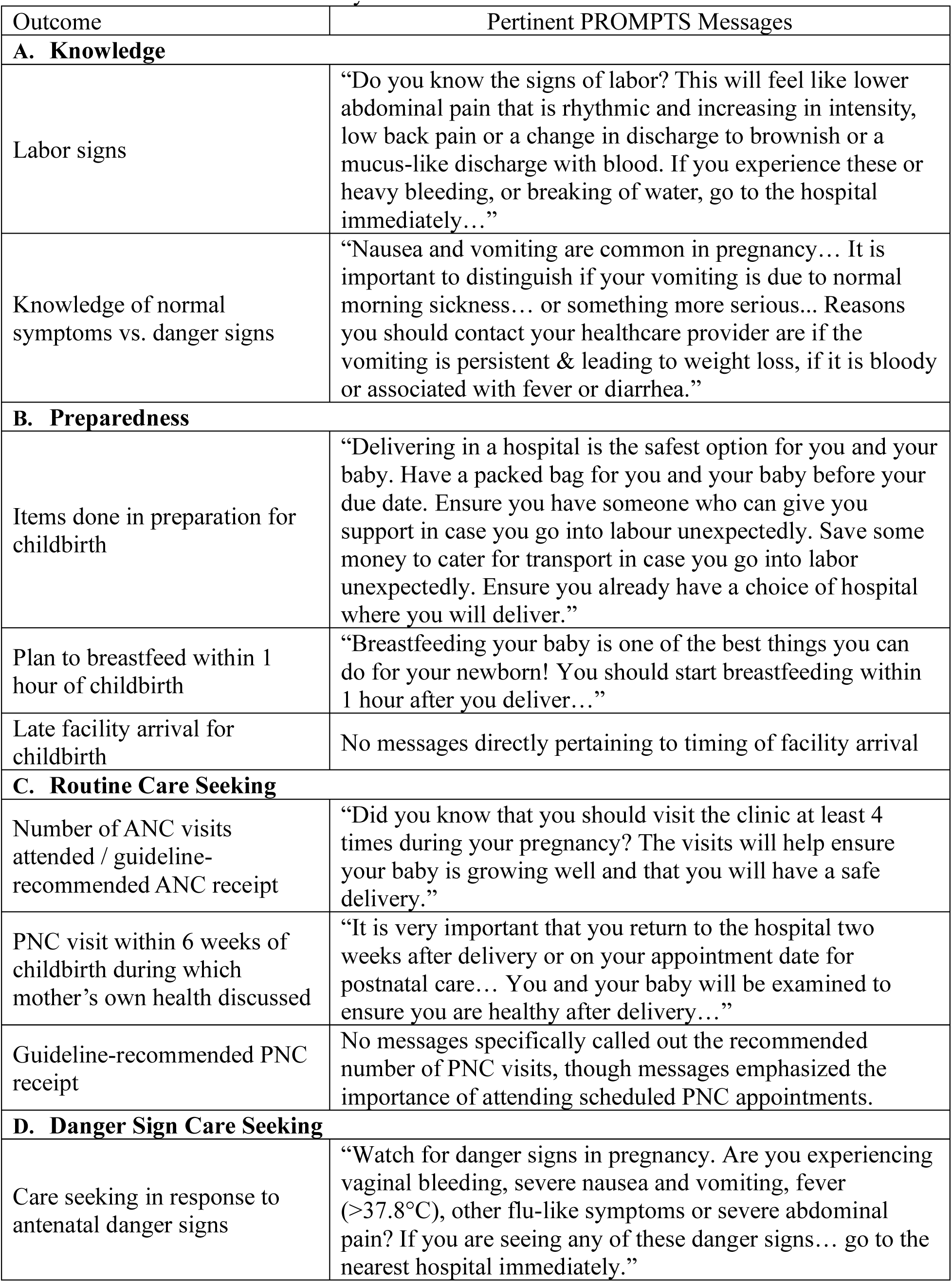
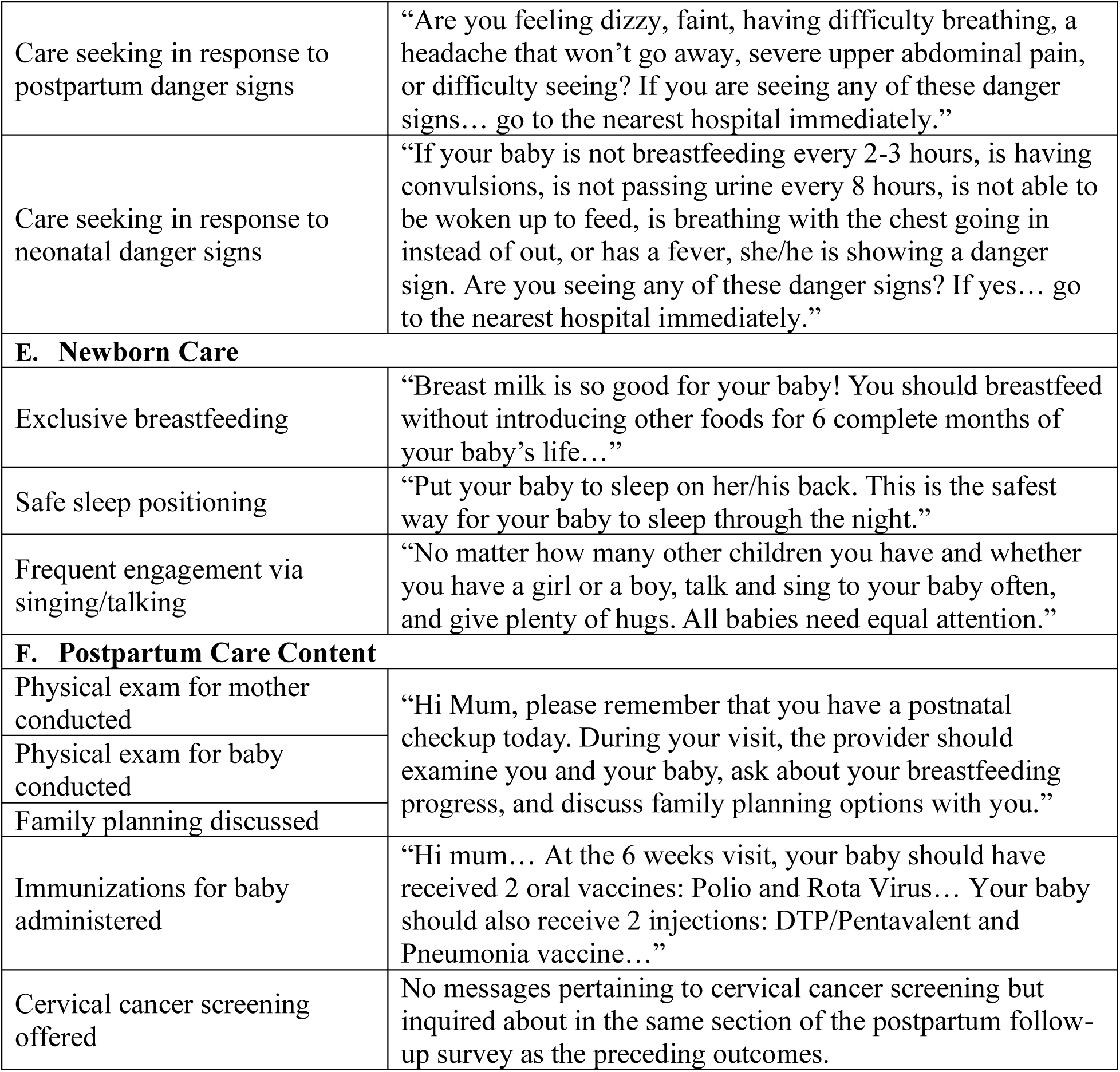
Outcome-Related Summary of SMS Content Delivered to PROMPTS Enrollees.

| Outcome | Pertinent PROMPTS Messages |
| --- | --- |
| **A. Knowledge** |  |
| Labor signs | “Do you know the signs of labor? This will feel like lower abdominal pain that is rhythmic and increasing in intensity, low back pain or a change in discharge to brownish or a mucus-like discharge with blood. If you experience these or heavy bleeding, or breaking of water, go to the hospital immediately...” |
| Knowledge of normal symptoms vs. danger signs | “Nausea and vomiting are common in pregnancy... It is important to distinguish if your vomiting is due to normal morning sickness... or something more serious... Reasons you should contact your healthcare provider are if the vomiting is persistent & leading to weight loss, if it is bloody or associated with fever or diarrhea.” |
| **B. Preparedness** |  |
| Items done in preparation for childbirth | “Delivering in a hospital is the safest option for you and your baby. Have a packed bag for you and your baby before your due date. Ensure you have someone who can give you support in case you go into labour unexpectedly. Save some money to cater for transport in case you go into labor unexpectedly. Ensure you already have a choice of hospital where you will deliver.” |
| Plan to breastfeed within 1 hour of childbirth | “Breastfeeding your baby is one of the best things you can do for your newborn! You should start breastfeeding within 1 hour after you deliver...” |
| Late facility arrival for childbirth | No messages directly pertaining to timing of facility arrival |
| **C. Routine Care Seeking** |  |
| Number of ANC visits attended / guideline-recommended ANC receipt | “Did you know that you should visit the clinic at least 4 times during your pregnancy? The visits will help ensure your baby is growing well and that you will have a safe delivery.” |
| PNC visit within 6 weeks of childbirth during which mother’s own health discussed | “It is very important that you return to the hospital two weeks after delivery or on your appointment date for postnatal care... You and your baby will be examined to ensure you are healthy after delivery...” |
| Guideline-recommended PNC receipt | No messages specifically called out the recommended number of PNC visits, though messages emphasized the importance of attending scheduled PNC appointments. |
| **D. Danger Sign Care Seeking** |  |
| Care seeking in response to antenatal danger signs | “Watch for danger signs in pregnancy. Are you experiencing vaginal bleeding, severe nausea and vomiting, fever (>37.8°C), other flu-like symptoms or severe abdominal pain? If you are seeing any of these danger signs... go to the nearest hospital immediately.” |
| Care seeking in response to postpartum danger signs | “Are you feeling dizzy, faint, having difficulty breathing, a headache that won’t go away, severe upper abdominal pain, or difficulty seeing? If you are seeing any of these danger signs... go to the nearest hospital immediately.” |
| Care seeking in response to neonatal danger signs | “If your baby is not breastfeeding every 2-3 hours, is having convulsions, is not passing urine every 8 hours, is not able to be woken up to feed, is breathing with the chest going in instead of out, or has a fever, she/he is showing a danger sign. Are you seeing any of these danger signs? If yes... go to the nearest hospital immediately.” |
| **E. Newborn Care** |  |
| Exclusive breastfeeding | “Breast milk is so good for your baby! You should breastfeed without introducing other foods for 6 complete months of your baby’s life...” |
| Safe sleep positioning | “Put your baby to sleep on her/his back. This is the safest way for your baby to sleep through the night.” |
| Frequent engagement via singing/talking | “No matter how many other children you have and whether you have a girl or a boy, talk and sing to your baby often, and give plenty of hugs. All babies need equal attention.” |
| **F. Postpartum Care Content** |  |
| Physical exam for mother conducted | “Hi Mum, please remember that you have a postnatal checkup today. During your visit, the provider should examine you and your baby, ask about your breastfeeding progress, and discuss family planning options with you.” |
| Physical exam for baby conducted |  |
| Family planning discussed |  |
| Immunizations for baby administered | “Hi mum... At the 6 weeks visit, your baby should have received 2 oral vaccines: Polio and Rota Virus... Your baby should also receive 2 injections: DTP/Pentavalent and Pneumonia vaccine...” |
| Cervical cancer screening offered | No messages pertaining to cervical cancer screening but inquired about in the same section of the postpartum follow-up survey as the preceding outcomes. |

Within the domain of patient knowledge, we constructed an index composed of four pre-registered outcomes: the shares of antenatal, postpartum, and neonatal danger sign knowledge questions answered correctly and the number of signs of labor listed, without prompting (see **Appendix S2** for details).

In terms of patient preparedness, we constructed an index composed of two pre-registered outcomes and one exploratory outcome. The first pre-registered outcome was the total number of items reported to have been completed, without prompting, in preparation for childbirth (see **Appendix S2** for details). The second was whether a participant had a plan to breastfeed within one hour of childbirth. Given PROMPTS’s objective to mitigate delays in care seeking, we also explored timeliness of facility arrival for childbirth, specifically whether a participant arrived late (i.e., within two hours of childbirth). We posited that this outcome may shift as a reflection of changes in preparedness.

Within the domain of routine care seeking, we constructed an index composed of two pre-registered outcomes and two exploratory outcomes. The pre-registered outcomes included the total number of antenatal care (ANC) visits attended and a binary indicator denoting receipt of postnatal care (PNC), within six weeks of childbirth, where one’s own health was discussed. In exploratory analyses of guideline-based care, we generated binary indicators denoting receipt of the guideline-recommended number of ANC and PNC visits, with guidelines recommending at least four ANC visits for women with uncomplicated pregnancies and at least two PNC visits following the first postpartum week.^39,40^

Within the domain of care seeking for danger signs, we constructed an index composed of three pre-registered outcomes, corresponding to each of three categories of severe danger signs: antenatal, postpartum, and neonatal. For each category, we assessed whether participants reported seeking medical advice or treatment from a healthcare provider in response to at least a single danger sign (see **Appendix S2** for details).

Within the domain of newborn care, we constructed an index composed of three pre-registered outcomes, namely binary indicators denoting whether participants reported exclusively breastfeeding, always putting their newborn to sleep through the night on their back, and singing or talking to their newborn many times during the preceding day.

Finally, within the domain of postpartum care content, we constructed an index composed of six exploratory outcomes: whether participants, during at least one PNC visit, reported that their health was discussed with a provider, that their provider conducted a physical exam for them, that their provider discussed family planning with them, that their provider offered them cervical cancer screening, that their provider conducted a physical exam for their newborn, and that their provider offered immunizations to their newborn.

### Statistical Analysis

Statistical analyses were conducted using Stata/MP 17.0 and R 4.2.3. The impacts of the intervention are presented as intention-to-treat (ITT) analyses, representing the impact of the offer of PROMPTS on outcomes. Impacts on individual-level binary and continuous outcome variables were estimated via ordinary least squares using linear (probability) models. With respect to the summary indices, the magnitudes of the impacts estimated can be interpreted as standard deviation changes in the respective domain in the treatment group compared to the control group. In our unadjusted regression specification, outcomes were regressed on a treatment indicator denoting whether an individual was recruited from a treatment facility and a stratification variable denoting the recruitment facility’s baseline normal vaginal birth volume tertile. We also estimated adjusted models in which we added a set of baseline individual- and facility-level covariates, including maternal age, gestational age, an indicator for first pregnancy, an indicator for secondary school attainment, an indicator for adequate prenatal care, an indicator for previous receipt of SMS messages from one’s county offering pregnancy advice, the recruitment facility level, and the recruitment facility county.

The addition of individual- and facility-level covariates was intended to improve precision of our average treatment effect estimates. Accordingly, covariates were chosen that we hypothesized would best account for residual variation in our outcomes of interest. Standard errors were clustered by enrollment facility, given the facility level randomization, and all results are presented as point estimates with 95% confidence intervals. Adjusted effect estimates are highlighted and discussed in the main text.

#### Sample Size and Power

The sample size of women who consented at baseline and the retention rate at postpartum follow-up (see **RESULTS**) both exceeded targets of 4,800 women and 75%, respectively. These targets were set to detect a minimum 6.5 percentage point (ppt) increase (11% relative increase) in the rate of postnatal care receipt within six weeks of childbirth, assuming a reference rate of 61% (based on internal data from Jacaranda Health), intracluster correlation of 0.01, type 1 error rate of 5%, and power of 80%. They were also set to detect a minimum 0.2 visit increase (5% relative increase) in the total number of ANC visits, assuming a reference mean of 4.12 and standard deviation (SD) of 1.73 (based on the 2014 Kenya Demographic and Health Survey).^41^

### Ethics and Safety Statement

This study was licensed by the National Commission for Science, Technology, and Innovation in Kenya (NACOSTI/P/21/13369) and approved by the institutional review board and ethics and scientific review committee of the Harvard T.H. Chan School of Public Health (IRB21-1013) and Amref Health Africa (P1047/2021). Written, electronic, or thumbprint-based informed consent was obtained from all participants. An emphasis on voluntary participation, participant confidentiality, and freedom to withdraw consent at any time was upheld throughout. **RESULTS**

### Sample Attrition and Characteristics

At baseline, 7,284 women were approached across the 40 study facilities, 6,139 of whom (3,140 in the treatment arm; 2,999 in the control arm) were eligible and consented to participate in the study (**Fig 2**). Only participants who were enrolled into the study prior to their 36^th^ week of gestation (N = 5,013) were contacted for the antenatal follow-up survey. Of those 5,013 women, 1,335 were ineligible due to their current pregnancy having already ended (with a live birth) when contacted. In these cases, these women were recontacted 7-8 weeks post-childbirth for the postpartum follow-up survey, unless their pregnancy had ended over six weeks ago, in which case they were immediately eligible for postpartum follow-up. 3,399 women (1,709 in the treatment arm; 1,690 in the control arm) ultimately completed the antenatal follow-up survey, representing an 8% loss to follow-up among those eligible (**Fig 2**). At postpartum follow-up, all 6,139 women who consented at baseline were contacted, and only 11 were ineligible due to being less than seven weeks postpartum. 5,509 women (2,833 in the treatment arm; 2,676 in the control arm) completed the survey, representing a 10% loss to follow-up among the eligible sample (**Fig 2**). At both antepartum and postpartum follow-up, the attrition rate did not significantly differ between study arms (**Table S1**).

Baseline characteristics of the study sample overall and by study arm are presented in **Table 2**. The average age of participants was 26 years, with 82% of the sample married or cohabitating and 65% having completed secondary school or higher. Study participants were at varying stages of pregnancy, with an average gestational age of 30 weeks and around 80% having received some form of prior antenatal care. Almost 90% of the participants had their own mobile phone, and 95% could read English or Kiswahili without difficulty. Approximately 22% reported having at least one high-risk condition (e.g., diabetes, pre-eclampsia, placenta previa) during the current pregnancy, as informed by a health provider, and among those with a prior pregnancy (66%), 43% reported history of either hypertension, pre-eclampsia, postpartum hemorrhage, preterm birth, stillbirth, neonatal death, or caesarean section. Finally, on average, participants correctly answered 68% of questions on a baseline knowledge assessment testing whether respondents would seek immediate medical care or watchfully wait in response to several potentially severe pregnancy symptoms. Sample characteristics were similar between study arms at baseline, as were the characteristics of those who completed the antenatal and postpartum follow-up surveys (**Tables 2, S2-S3**).

**Table 2:** Baseline Characteristics Among Eligible and Consented Cohort of Pregnant Women Recruited During ANC Visits to Study Facilities.

| Baseline Characteristic | Overall<br>(N = 6,139) | Control Arm<br>(N = 2,999) | Treated Arm<br>(N = 3,140) |
| --- | --- | --- | --- |
| Age (years) | 26.15 (5.89) | 25.95 (5.94) | 26.35 (5.84) |
| Secondary school education or higher | 64.8% | 63.6% | 66.1% |
| Read Kiswahili or English without difficulty | 95.2% | 94.8% | 95.6% |
| Married or living together | 82.1% | 81.3% | 82.9% |
| Size of household | 3.81 (1.99) | 4.01 (2.13) | 3.62 (1.84) |
| Own land | 42.4% | 47.1% | 37.8% |
| Improved source of drinking water | 70.1% | 69.3% | 70.8% |
| Improved sanitation facility | 97.7% | 97.2% | 98.3% |
| Mode of travel to hospital: motor vehicle | 69.3% | 69.6% | 69.0% |
| Time to travel from home to health facility (minutes) | 23.92 (18.54) | 24.64 (18.64) | 23.23 (18.43) |
| Work for pay in last week | 22.6% | 22.3% | 23.0% |
| Easy access to Ksh 2000 if treatment for illness needed in household | 27.5% | 27.3% | 27.6% |
| Own mobile phone | 89.6% | 88.0% | 91.1% |
| Use mobile phone to send text messages frequently/daily | 36.7% | 33.5% | 39.8% |
| Previous receipt of text message offering pregnancy advice from county | 5.0% | 4.0% | 6.0% |
| Gestational age (weeks) | 29.69 (6.14) | 29.61 (6.16) | 29.76 (6.12) |
| Any prior prenatal care visit | 80.9% | 80.7% | 81.1% |
| # prenatal care visits for current pregnancy | 1.93 (1.52) | 1.90 (1.48) | 1.97 (1.55) |
| Fraction of knowledge questions answered correctly | 0.68 (0.18) | 0.67 (0.18) | 0.69 (0.18) |
| High-risk pregnancy | 21.8% | 23.6% | 20.0% |
| # of total pregnancies, including current pregnancy | 2.37 (1.47) | 2.42 (1.51) | 2.32 (1.42) |
| Previous high-risk pregnancy | 42.6% | 41.1% | 44.1% |
| PHQ-2 score | 1.57 (1.60) | 1.51 (1.58) | 1.63 (1.61) |

Abbreviations: ANC, antenatal care; PHQ-2, Patient Health Questionnaire-2
Continuous variables summarized by sample mean and standard deviation: mean (SD); binary variables summarized by sample mean as a %.

### Intervention Fidelity

Fidelity and uptake of the PROMPTS platform is summarized in **Table 3**, which reveals that among participants from treatment facilities, over 85% reported receipt of messages offering pregnancy advice, while less than 10% of participants from control facilities reported such receipt. Thus, recruitment from treatment facilities was associated with an ∼75ppt increase in the take-up of PROMPTS.

**Table 3:** Intervention Fidelity Ascertained at Antenatal Follow-Up.

|  | Control Mean | Treatment Mean | Unadjusted Treatment Effect <sup>a</sup> | Adjusted Treatment Effect <sup>b</sup> |
| --- | --- | --- | --- | --- |
| Share of participants who reported receipt of any messages from their county offering pregnancy advice <sup>c</sup> | 0.09 | 0.85 | 0.76 **<br>95% CI: (0.73, 0.79)<br>P < 0.01 | 0.76 **<br>95% CI: (0.73, 0.78)<br>P < 0.01 |

\* $p < 0.05$ \*\* $p < 0.01$
<sup>a</sup> The unadjusted model only includes covariates pertinent to the randomization procedure (e.g., recruitment facility's baseline normal vaginal birth volume tertile).
<sup>b</sup> The adjusted model adds baseline individual- and recruitment-facility-level covariates, including maternal age, gestational age, an indicator for first pregnancy, an indicator for secondary school attainment, an indicator for adequate prenatal care, an indicator for previous receipt of SMS messages from one's county offering pregnancy advice, facility level, and facility county.
<sup>c</sup> Ascertained via phone surveys administered by study enumerators at antenatal follow-up.

### Intervention Impact

Women in the treatment arm had a 0.08 SD (95% CI: 0.03, 0.12) higher knowledge index compared to those in the control arm, with impacts concentrated in the antenatal period (**Table 4**). Specifically, those in the treatment arm had a 3.6ppt (95% CI: 1.9, 5.4) higher antenatal danger sign knowledge score and could list 0.24 (95% CI: 0.16, 0.32) more signs of labor than participants in the control arm. No statistically significant differences in the postpartum or neonatal danger sign knowledge indices were found.

**Table 4:**
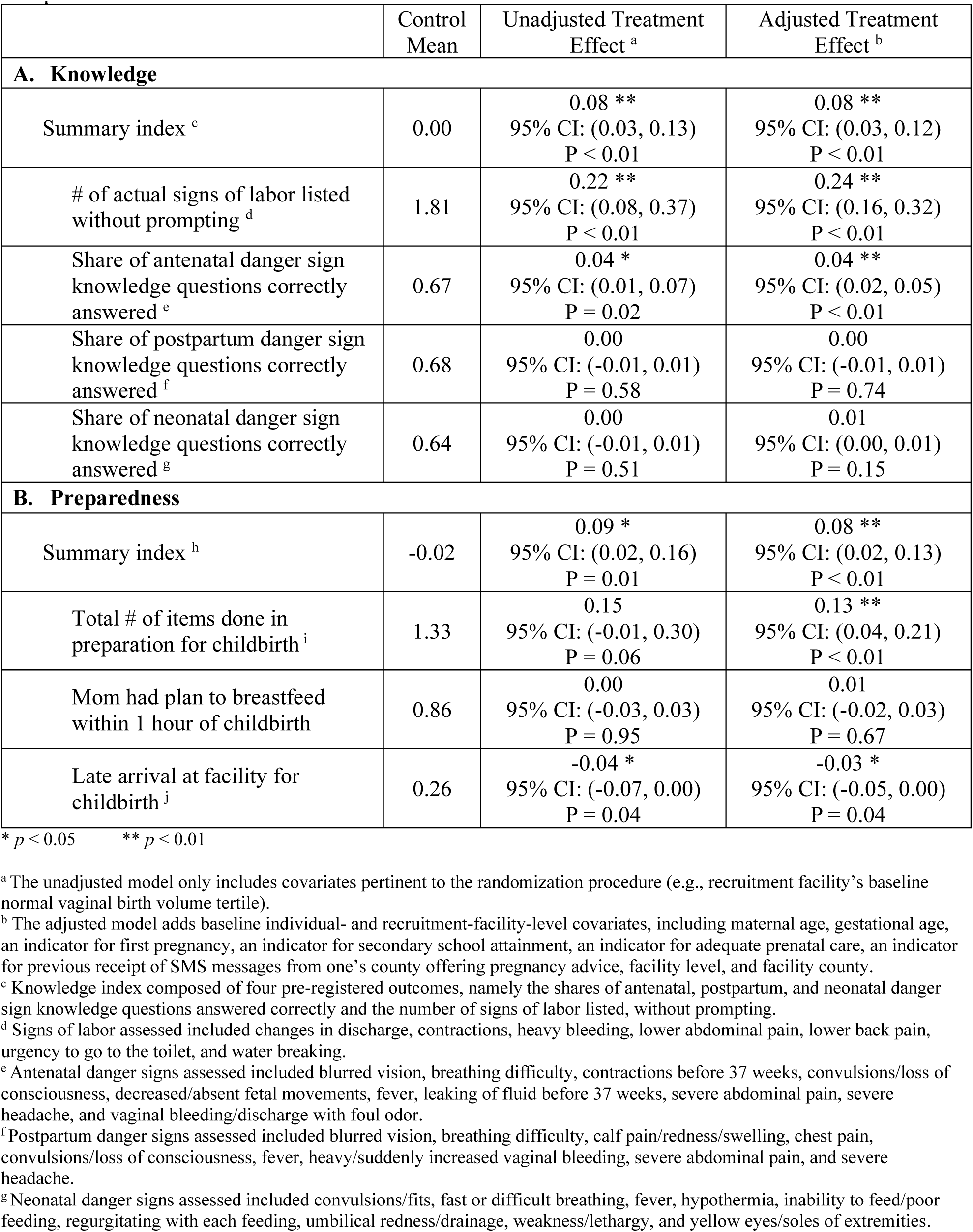
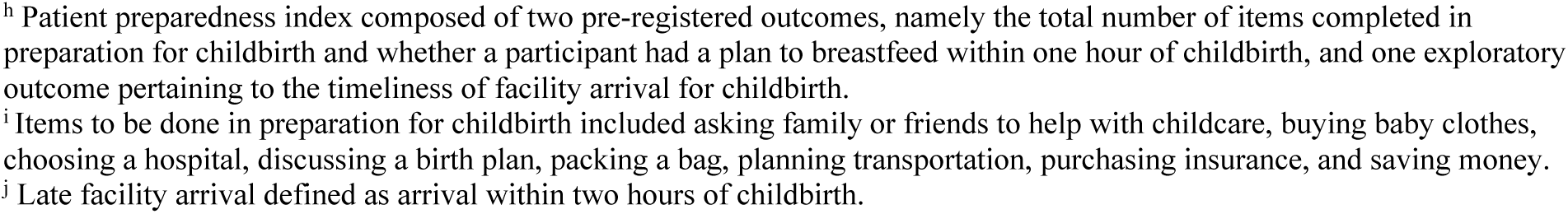
Impact of Study Intervention on Knowledge and Preparedness Across the Pregnancy-Postpartum Care Continuum.

|  | Control Mean | Unadjusted Treatment Effect <sup>a</sup> | Adjusted Treatment Effect <sup>b</sup> |
| --- | --- | --- | --- |
| **A. Knowledge** |  |  |  |
| Summary index <sup>c</sup> | 0.00 | 0.08 **<br>95% CI: (0.03, 0.13)<br>P < 0.01 | 0.08 **<br>95% CI: (0.03, 0.12)<br>P < 0.01 |
| # of actual signs of labor listed without prompting <sup>d</sup> | 1.81 | 0.22 **<br>95% CI: (0.08, 0.37)<br>P < 0.01 | 0.24 **<br>95% CI: (0.16, 0.32)<br>P < 0.01 |
| Share of antenatal danger sign knowledge questions correctly answered <sup>e</sup> | 0.67 | 0.04 *<br>95% CI: (0.01, 0.07)<br>P = 0.02 | 0.04 **<br>95% CI: (0.02, 0.05)<br>P < 0.01 |
| Share of postpartum danger sign knowledge questions correctly answered <sup>f</sup> | 0.68 | 0.00<br>95% CI: (-0.01, 0.01)<br>P = 0.58 | 0.00<br>95% CI: (-0.01, 0.01)<br>P = 0.74 |
| Share of neonatal danger sign knowledge questions correctly answered <sup>g</sup> | 0.64 | 0.00<br>95% CI: (-0.01, 0.01)<br>P = 0.51 | 0.01<br>95% CI: (0.00, 0.01)<br>P = 0.15 |
| **B. Preparedness** |  |  |  |
| Summary index <sup>h</sup> | -0.02 | 0.09 *<br>95% CI: (0.02, 0.16)<br>P = 0.01 | 0.08 **<br>95% CI: (0.02, 0.13)<br>P < 0.01 |
| Total # of items done in preparation for childbirth <sup>i</sup> | 1.33 | 0.15<br>95% CI: (-0.01, 0.30)<br>P = 0.06 | 0.13 **<br>95% CI: (0.04, 0.21)<br>P < 0.01 |
| Mom had plan to breastfeed within 1 hour of childbirth | 0.86 | 0.00<br>95% CI: (-0.03, 0.03)<br>P = 0.95 | 0.01<br>95% CI: (-0.02, 0.03)<br>P = 0.67 |
| Late arrival at facility for childbirth <sup>j</sup> | 0.26 | -0.04 *<br>95% CI: (-0.07, 0.00)<br>P = 0.04 | -0.03 *<br>95% CI: (-0.05, 0.00)<br>P = 0.04 |

\* $p < 0.05$ \*\* $p < 0.01$
<sup>a</sup> The unadjusted model only includes covariates pertinent to the randomization procedure (e.g., recruitment facility's baseline normal vaginal birth volume tertile).
<sup>b</sup> The adjusted model adds baseline individual- and recruitment-facility-level covariates, including maternal age, gestational age, an indicator for first pregnancy, an indicator for secondary school attainment, an indicator for adequate prenatal care, an indicator for previous receipt of SMS messages from one's county offering pregnancy advice, facility level, and facility county.
<sup>c</sup> Knowledge index composed of four pre-registered outcomes, namely the shares of antenatal, postpartum, and neonatal danger sign knowledge questions answered correctly and the number of signs of labor listed, without prompting.
<sup>d</sup> Signs of labor assessed included changes in discharge, contractions, heavy bleeding, lower abdominal pain, lower back pain, urgency to go to the toilet, and water breaking.
<sup>e</sup> Antenatal danger signs assessed included blurred vision, breathing difficulty, contractions before 37 weeks, convulsions/loss of consciousness, decreased/absent fetal movements, fever, leaking of fluid before 37 weeks, severe abdominal pain, severe headache, and vaginal bleeding/discharge with foul odor.
<sup>f</sup> Postpartum danger signs assessed included blurred vision, breathing difficulty, calf pain/redness/swelling, chest pain, convulsions/loss of consciousness, fever, heavy/suddenly increased vaginal bleeding, severe abdominal pain, and severe headache.
<sup>g</sup> Neonatal danger signs assessed included convulsions/fits, fast or difficult breathing, fever, hypothermia, inability to feed/poor feeding, regurgitating with each feeding, umbilical redness/drainage, weakness/lethargy, and yellow eyes/soles of extremities.
<sup>h</sup> Patient preparedness index composed of two pre-registered outcomes, namely the total number of items completed in preparation for childbirth and whether a participant had a plan to breastfeed within one hour of childbirth, and one exploratory outcome pertaining to the timeliness of facility arrival for childbirth. <sup>i</sup> Items to be done in preparation for childbirth included asking family or friends to help with childcare, buying baby clothes, choosing a hospital, discussing a birth plan, packing a bag, planning transportation, purchasing insurance, and saving money. <sup>j</sup> Late facility arrival defined as arrival within two hours of childbirth.

Women in the treatment arm also had a 0.08 SD (95% CI: 0.02, 0.13) higher preparedness index compared to those in the control arm (**Table 4**). This included completing 0.13 (95% CI: 0.04, 0.21) more items in preparation for childbirth, with an over two-fold (1.9ppt; 95% CI: 0.7, 3.1) increase in the share of women purchasing health insurance. No statistically significant difference in women’s plans to breastfeed within one hour of childbirth was found. Meanwhile, women in the treatment arm were 2.8ppt (95% CI: -5.4, -0.2) less likely to arrive at a facility within two hours of childbirth than women in the control arm (**Fig 4**).

**Fig 4:**
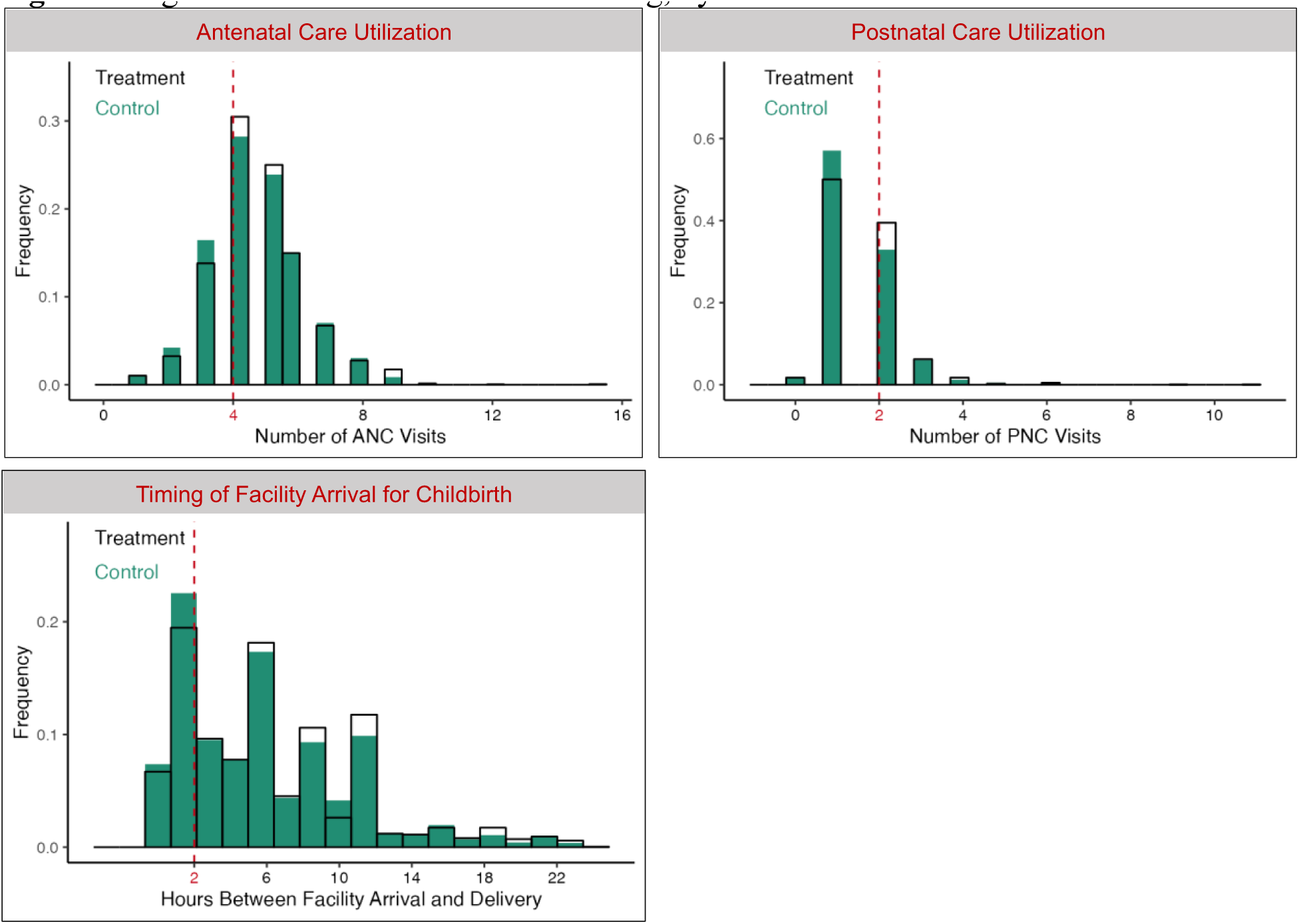
Histograms of Care Utilization and Timing, by Treatment Arm. This figure displays the increase in volume of antenatal and postnatal care visits among participants recruited from treatment facilities just at or above the guideline-based thresholds of ≥4 ANC visits throughout pregnancy and ≥2 PNC visits during the first six weeks postpartum, following the first postpartum week. It also displays visual evidence of an increase in the timing between facility arrival and childbirth (i.e., reduction in late facility arrival) for participants recruited from treatment facilities.

With respect to routine care seeking, women in the treatment arm had a 0.10 SD (95% CI: 0.05, 0.16) higher summary index compared to those in the control arm (**Table 5**). For antenatal care, the share of women receiving at least the guideline-recommended number of four ANC visits was 3.1ppt (95% CI: 0.4, 5.7) higher in the treatment arm, though no statistically significant difference in the total number of ANC visits was found. As for postnatal care, there was a 4.8ppt (95% CI: 0.7, 9.0) increase in the share of women who attended at least one PNC visit within six weeks of childbirth during which their own health was discussed, as well as a 7.4ppt (95% CI: 3.5, 11.3) increase in the share of women receiving at least the guideline-recommended number of PNC visits. The latter corresponded to an 18% relative increase in the treatment arm. Of note, increases in ANC and PNC utilization appeared to be concentrated around the guideline-recommended thresholds for antenatal and postnatal care (**Fig 4**).

**Table 5:** Impact of Study Intervention on Care Seeking, Behavior, and Content Across the Pregnancy-Postpartum Care Continuum.

|  | Control<br>Mean | Unadjusted Treatment<br>Effect <sup>a</sup> | Adjusted Treatment<br>Effect <sup>b</sup> |
| --- | --- | --- | --- |
| **A. Routine Care Seeking** |  |  |  |
| Summary index <sup>c</sup> | 0.00 | 0.10<br>95% CI: (-0.01, 0.21)<br>P = 0.08 | 0.10 **<br>95% CI: (0.05, 0.16)<br>P < 0.01 |
| Total # of ANC visits attended | 4.65 | 0.08<br>95% CI: (-0.18, 0.34)<br>P = 0.55 | 0.10<br>95% CI: (-0.04, 0.23)<br>P = 0.16 |
| At least 1 PNC visit attended by mother within 6 weeks of childbirth during which her own health was discussed | 0.64 | 0.05<br>95% CI: (-0.02, 0.12)<br>P = 0.20 | 0.05 *<br>95% CI: (0.01, 0.09)<br>P = 0.02 |
| Received at least the guideline-recommended # of ANC visits <sup>d</sup> | 0.78 | 0.04<br>95% CI: (-0.01, 0.08)<br>P = 0.15 | 0.03 *<br>95% CI: (0.00, 0.06)<br>P = 0.02 |
| Received at least the guideline-recommended # of PNC visits <sup>e</sup> | 0.41 | 0.07<br>95% CI: (-0.01, 0.14)<br>P = 0.08 | 0.07 **<br>95% CI: (0.04, 0.11)<br>P < 0.01 |
| **B. Danger Sign Care Seeking** |  |  |  |
| Summary index <sup>f</sup> | 0.02 | 0.03<br>95% CI: (-0.02, 0.09)<br>P = 0.27 | 0.04<br>95% CI: (-0.01, 0.08)<br>P = 0.10 |
| Medical care sought for mother in response to ≥1 antenatal danger sign <sup>g</sup> | 0.54 | 0.02<br>95% CI: (-0.03, 0.07)<br>P = 0.34 | 0.03<br>95% CI: (0.00, 0.06)<br>P = 0.08 |
| Medical care sought for mother in response to ≥1 postpartum danger sign <sup>h</sup> | 0.14 | 0.02<br>95% CI: (0.00, 0.04)<br>P = 0.08 | 0.02 *<br>95% CI: (0.00, 0.04)<br>P = 0.02 |
| Medical care sought for baby in response to ≥1 neonatal danger sign <sup>i</sup> | 0.22 | 0.01<br>95% CI: (-0.02, 0.03)<br>P = 0.54 | 0.01<br>95% CI: (-0.01, 0.03)<br>P = 0.36 |
| **C. Newborn Care** |  |  |  |
| Summary index <sup>j</sup> | 0.00 | 0.10 **<br>95% CI: (0.06, 0.14)<br>P < 0.01 | 0.09 **<br>95% CI: (0.07, 0.12)<br>P < 0.01 |
| Newborn exclusively breastfed by mother | 0.93 | 0.01<br>95% CI: (-0.01, 0.03)<br>P = 0.26 | 0.01<br>95% CI: (0.00, 0.02)<br>P = 0.14 |
| Newborn always put to sleep through the night on their back | 0.03 | 0.02 *<br>95% CI: (0.00, 0.03)<br>P = 0.04 | 0.02 **<br>95% CI: (0.01, 0.03)<br>P < 0.01 |
| Newborn sung/talked to by mother many times over past 24 hours | 0.82 | 0.06 **<br>95% CI: (0.03, 0.09)<br>P < 0.01 | 0.05 **<br>95% CI: (0.03, 0.07)<br>P < 0.01 |
| **D. Postpartum Care Content** |  |  |  |
| Summary index <sup>k</sup> | 0.00 | 0.06 | 0.06 * |
|  |  | 95% CI: (-0.01, 0.14)<br>P = 0.11 | 95% CI: (0.01, 0.12)<br>P = 0.02 |
| Mother's health discussed with a provider during at least one PNC visit | 0.63 | 0.05<br>95% CI: (-0.03, 0.12)<br>P = 0.21 | 0.05 *<br>95% CI: (0.00, 0.09)<br>P = 0.03 |
| Provider conducted physical exam for mother during at least one PNC visit | 0.54 | 0.07<br>95% CI: (-0.01, 0.15)<br>P = 0.10 | 0.06 *<br>95% CI: (0.01, 0.11)<br>P = 0.01 |
| Provider discussed family planning with mother during at least one PNC visit | 0.53 | 0.08<br>95% CI: (-0.01, 0.16)<br>P = 0.07 | 0.07 *<br>95% CI: (0.01, 0.14)<br>P = 0.02 |
| Provider offered mother cervical cancer screening during at least one PNC visit | 0.11 | 0.00<br>95% CI: (-0.03, 0.02)<br>P = 0.90 | 0.00<br>95% CI: (-0.02, 0.03)<br>P = 0.72 |
| Provider conducted physical exam for baby during at least one PNC visit | 0.95 | 0.01<br>95% CI: (0.00, 0.03)<br>P = 0.08 | 0.01 *<br>95% CI: (0.00, 0.02)<br>P = 0.02 |
| Provider provided immunization for baby during at least one PNC visit | 0.98 | 0.00<br>95% CI: (-0.01, 0.01)<br>P = 0.72 | 0.00<br>95% CI: (-0.01, 0.00)<br>P = 0.48 |

\* $p < 0.05$ \*\* $p < 0.01$
Abbreviations: ANC, antenatal care; PNC, postnatal/postpartum care
<sup>a</sup> The unadjusted model only includes covariates pertinent to the randomization procedure (e.g., recruitment facility's baseline normal vaginal birth volume tertile).
<sup>b</sup> The adjusted model adds baseline individual- and recruitment-facility-level covariates, including maternal age, gestational age, an indicator for first pregnancy, an indicator for secondary school attainment, an indicator for adequate prenatal care, an indicator for previous receipt of SMS messages from one's county offering pregnancy advice, facility level, and facility county.
<sup>c</sup> Routine care seeking index composed of two pre-registered outcomes, namely the total number of ANC visits attended and whether a participant received postnatal care with their own health discussed at least once within six weeks of childbirth, and two exploratory outcomes pertaining to receipt of the guideline-recommended quantity of routine antenatal and postnatal care.
<sup>d</sup> Guidelines recommend at least four ANC visits across pregnancy for women with uncomplicated pregnancies and more otherwise.
<sup>e</sup> Following the first postpartum week, WHO guidelines recommend at least two additional PNC visits, between weeks 1-2 and during week 6 postpartum.
<sup>f</sup> Danger sign care seeking index composed of three pre-registered outcomes, corresponding to each of three categories of severe danger signs: antenatal, postpartum, and neonatal. For each category, we assessed whether participants reported seeking medical advice or treatment from a healthcare provider in response to at least a single danger sign.
<sup>g</sup> Antenatal danger signs assessed included blurred vision, breathing difficulty, contractions before 37 weeks, convulsions/loss of consciousness, decreased/absent fetal movements, fever, leaking of fluid before 37 weeks, severe abdominal pain, severe headache, and vaginal bleeding/discharge with foul odor.
<sup>h</sup> Postpartum danger signs assessed included blurred vision, breathing difficulty, calf pain/redness/swelling, chest pain, convulsions/loss of consciousness, fever, heavy/suddenly increased vaginal bleeding, severe abdominal pain, and severe headache.
<sup>i</sup> Neonatal danger signs assessed included convulsions/fits, fast or difficult breathing, fever, hypothermia, inability to feed/poor feeding, regurgitating with each feeding, umbilical redness/drainage, weakness/lethargy, and yellow eyes/soles of extremities.
<sup>j</sup> Newborn care index composed of three pre-registered outcomes, namely indicators denoting whether participants reported exclusively breastfeeding, always putting their newborn to sleep through the night on their back, and singing or talking to their newborn many times during the preceding day.
<sup>k</sup> Postpartum care content index composed of six exploratory outcomes, namely indicators denoting whether, during at least once PNC visit, participants reported that their health was discussed with a provider, that their provider conducted a physical exam for them, that their provider discussed family planning with them, that their provider offered them cervical cancer screening, that their provider conducted a physical exam for their newborn, and that their provider offered immunizations to their newborn.

Turning to care seeking in response to danger signs, no statistically significant difference across the treatment and control groups was found in the summary index (**Table 5**). However, the treatment group did have a 2.1ppt (95% CI: 0.4, 3.7) higher rate of care seeking for postpartum maternal danger signs than the control group.

The intervention led to a 0.09 SD (95% CI: 0.07, 0.12) higher newborn care index (**Table 5**), driven by improvements in newborn sleep positioning and engagement. Specifically, the share of mothers in the treatment group who reported always putting their newborn to sleep at night on their back was 1.9ppt (95% CI: 1.0, 2.8) higher than in the control group and the share of mothers who reported frequently engaging with their newborn through singing and talking was 5.2ppt (95% CI: 3.2, 7.2) higher. No statistically significant difference in the rate of exclusive breastfeeding was found, though the control group rate was already quite high at 93%.

Finally, the intervention led to a 0.06 SD (95% CI: 0.01, 0.12) higher postpartum care content index (**Table 5**). As depicted in **Fig 5**, improvements in the overall domain were driven by impacts on the content of care that mothers – as opposed to their newborns – received during at least one postpartum checkup. These chiefly included discussions of mothers’ own health, counseling on family planning, and receipt of a physical examination, which had relative increases of 8%, 11%, and 13%, respectively, in the treatment arm compared to the control arm. On the other hand, no meaningful differences were found in the frequency of providers conducting newborn physical exams or administering newborn vaccines, with rates of these practices already quite high in the control group (95% and 98%, respectively). No difference across groups was found in the rate of mothers being offered cervical cancer screening either.

**Fig 5:**
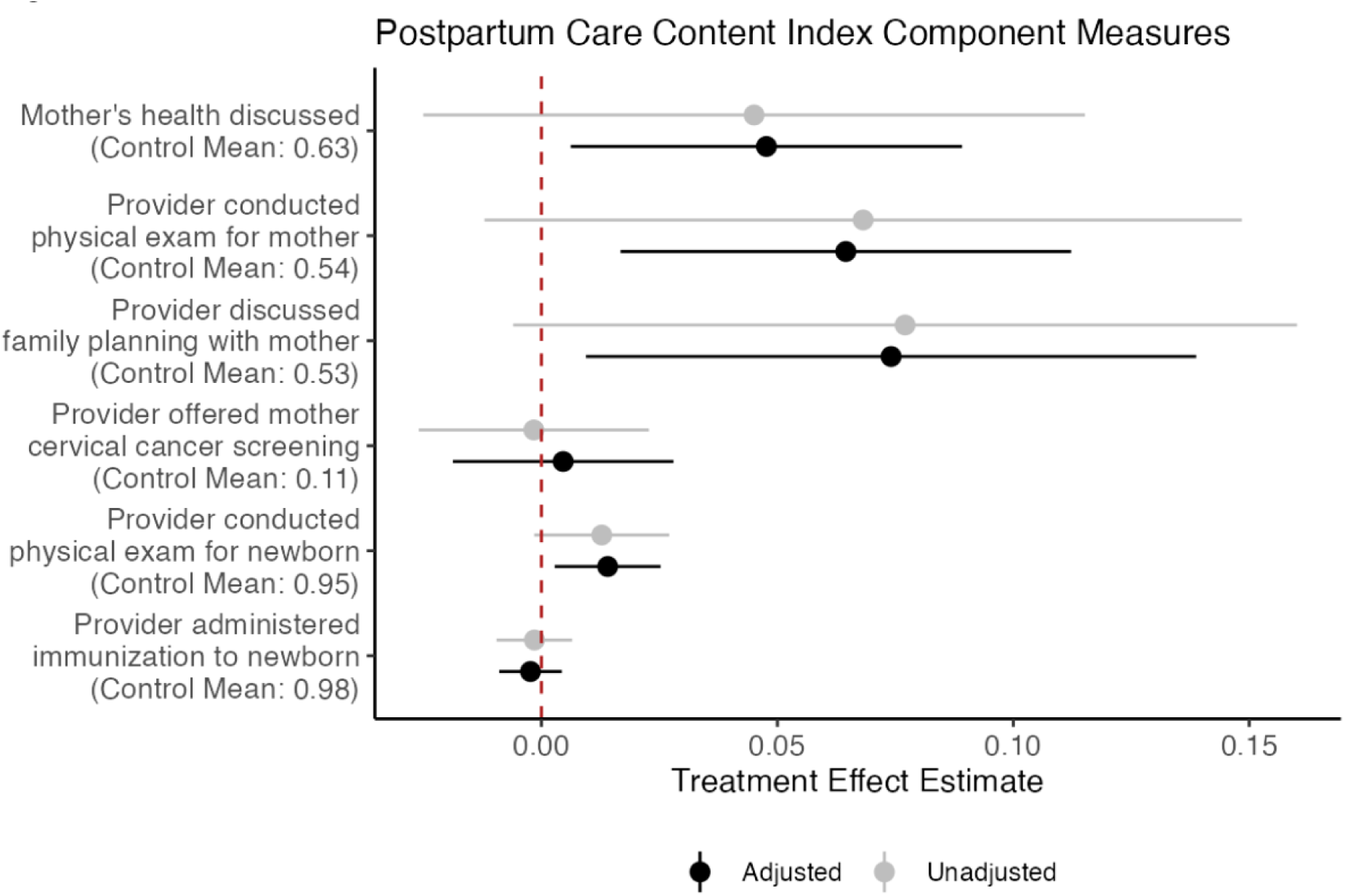
Impact of PROMPTS on the Content of Care Received by Mothers and Their Newborns During PNC Visits. This figure depicts adjusted and unadjusted treatment effects for the six component measures incorporated in the postpartum care content index. The figure reveals that overall improvements in the domain were driven by improvements in care that mothers received (e.g., more frequent discussions of their own health and discussions of family planning).

## DISCUSSION

We found that the PROMPTS digital health platform led to a range of improvements across the pregnancy-postpartum care continuum, including in participants’ knowledge, preparedness, routine and danger sign care seeking, newborn care, and postpartum care content. Although the standardized effect sizes that we identify for each index are, in isolation, modest, we find directionally consistent improvements across all domains. Moreover, given the real-world scale of the intervention, even such modest changes can be clinically significant.

Our results revealed particular gains in the postpartum setting, with an emphasis on mothers in addition to their newborns. Rates of postpartum care increased, both for routine checkups and maternal danger signs, with a nearly 20% rise in the share of women receiving at least the guideline-recommended number of PNC visits. Moreover, the content of postpartum checkups featured a renewed focus on mothers, with an over 10% increase in rates of family planning counseling and receipt of a physical examination. Of note, maternal care content measures were substantially lower than neonatal measures in the control group, offering an increased opportunity for PROMPTS to influence care for mothers. While other effects pertaining to knowledge, preparedness, and care seeking were more modest, the increases may still be meaningful. For one, Jacaranda Health estimates that PROMPTS costs a mere 74 cents per participant for their lifetime on the platform, and PROMPTS has been implemented at remarkable scale, with over two million individuals enrolled as of November 2023.^36^

It is also worth noting that our estimates may constitute a lower bound of PROMPTS’s potential impact. Specifically, our ITT estimates capture the impact of the offer of PROMPTS and do not account for take-up of the platform. Accounting for relative take-up (i.e., scaling our estimates by 1/[treatment group take-up – control group take-up] ≈ 1/0.76 ≈ 31%) may better isolate the impact of adopting the platform. On the other hand, imperfect adoption is to be expected with any intervention, and perhaps the 76ppt higher platform take-up that we identify is a reasonable indication of real-world adoption. In fact, routinely collected administrative data from Jacaranda Health suggest that only ¾ of enrollees actively engage with PROMPTS (e.g., by responding to questions administered on the platform or submitting questions of their own).

With respect to our estimated effects, the improvements in knowledge we identified were concentrated in the prenatal as opposed to postnatal period, with a 5% increase in antenatal danger sign knowledge scores and 13% increase in the number of labor signs listed. This could be attributed to participants’ increased attention to PROMPTS message content in the initial phases of enrollment and decreased attention amidst the heightened demands of postpartum life. Moreover, the ostensible fact that improvements in antenatal knowledge did not translate to as substantial gains in care seeking could be explained by already high levels of antenatal care in the control group. Consistent with Kenya’s historic emphasis on maternal care during this period, nearly 80% of women in the control group attended at least four ANC visits, with over 50% seeking care for at least one antenatal danger sign.^8,10,11^ Nevertheless, our findings are consistent with previous studies, which have highlighted the role of mobile health in patient education, including a study in India that found that SMS messages led to improvements in knowledge, particularly for mothers in the antenatal setting.^30,42^ Moreover, though PROMPTS may overcome a number of hurdles that health system level approaches to improving knowledge have faced (e.g., loss of in-person follow-up postpartum, inadequate CHW coverage), neither have been as successful at knowledge improvement in the postpartum period.^43,44,45^

Prior qualitative work has also suggested a role for SMS messages in promoting increased patient engagement, healthy behaviors, and timely care seeking during pregnancy.^46^ Relatedly, we observed a nearly 10% increase in the number of items that women in the treatment arm completed in preparation for childbirth. To the extent that such improvements translated to enhanced timeliness of delivery care, we also observed a potentially associated decrease in the rate of late facility arrival for childbirth. Our results on preparedness behaviors align with literature on a specific class of interventions geared towards improving birth preparedness and complication readiness (BPCR). Such interventions are premised on the notion that improvements in preparedness (e.g., saving money, arranging transportation) will facilitate timely use of care. While successful, BPCR interventions have historically operated at the health system level, through in-person counseling, or via community mobilization efforts.^47^

With respect to antenatal and postpartum care seeking, our results are situated in an extensive literature demonstrating the effectiveness of mobile health tools and other patient level interventions, particularly for increasing antenatal care attendance.^30,31,32^ While prior studies across sub-Saharan Africa have identified larger effect sizes than what we estimate (10ppt increases in ANC attendance, 20ppt increase in PNC attendance), those studies were either smaller in scale, conducted in a single site, and/or based in settings over a decade ago where baseline rates of formal care seeking were lower. ^48,49,50^ Moreover, though prior work on the cost-effectiveness of such strategies is limited, our findings align in magnitude with prior health system level initiatives that have been deemed cost-effective in increasing rates of guideline-recommended antenatal care.^28,51^ PROMPTS also impacts a broader array of domains in a more targeted fashion than initiatives with a lower cost profile (e.g., national media campaigns).^52^

As for danger signs, prior work has suggested difficulty in improving care seeking for maternal illnesses in the absence of comprehensive health system level support, including CHW home visits, counseling, and danger sign recognition.^27^ By contrast, we found that PROMPTS led to elevated care seeking for maternal danger signs, particularly in the postpartum period.

However, we found no such effect on care seeking for neonatal danger signs, which prior health system level approaches have demonstrated modest success in improving.

Our results on postpartum care content and newborn care are also situated in an emerging literature focused on the postnatal setting. Prior work in sub-Saharan Africa on facility-based provider trainings found similar increases in the rate of family planning counseling as our study.^43^ While encouraging, other work highlights limitations in the extent of content covered through such health system level strategies, particularly in overburdened delivery settings, and acknowledges the potential of post-discharge SMS outreach.^43,45^ Accordingly, prior evaluations of mobile health interventions have demonstrated sizable improvements in rates of exclusive breastfeeding.^32^ While we identify no such effect, rates of exclusive breastfeeding were quite high in our control sample (93%), unlike in the other settings evaluated. Indeed, for behaviors with lower baseline adoption in our study (e.g., newborn sleep positioning and engagement), we find notable improvements. While the cost-effectiveness of interventions to improve postnatal care in lower-income settings remains a gap in the literature, our findings suggest a broad role for low-cost mobile health tools like PROMPTS to advance both postpartum and newborn care.^28^

### Strengths and Limitations

The key strengths of our study were its size and breadth: we recruited and followed over 6,000 individuals across 40 health facilities in eight Kenyan counties, and rather than focus on a narrow set of metrics, we examined several domains across the pregnancy-postpartum care continuum. Moreover, we evaluated a cluster RCT to obtain causal impact estimates.

Nonetheless, our study also had limitations. First, we examined a substantial number of outcomes, which increased the risk of false positive conclusions. However, we mitigated this risk by organizing our outcomes into domains and generating summary indices. Second, all outcomes were self-reported, raising the risk of recall bias or social-desirability bias. Third, the interaction of PROMPTS with Jacaranda Health’s complementary mentorship program could have plausibly influenced outcomes in the childbirth and postpartum settings. Specifically, provider-facing trainings could have influenced patients’ childbirth experiences and the extent to which they felt empowered to return for postpartum care. However, it is important to note that provider trainings emphasized care for relatively rare emergencies, potentially alleviating this concern. Fourth, the COVID-19 pandemic may have dampened the impact of PROMPTS on care seeking behavior.

Nonetheless, by March 2021, over half a year before baseline data collection, 96% of households in Kenya were able to access health services as before the pandemic, and by May 2021, major lockdowns had been lifted.^53^ Finally, our study was not sufficiently powered to detect effects on major health outcomes, such as maternal and perinatal mortality.

### Implications and Future Directions

To our knowledge, our paper is the first to demonstrate the impact of an at-scale digital health platform across the pregnancy-postpartum care continuum in sub-Saharan Africa. While we identified several effects across the continuum, we found notable advances in the postpartum setting, for both mothers and babies. In almost all cases, the estimated effects were for outcomes evaluated based on PROMPTS content sent to enrollees (see **Table 1**), with null effects for those without corresponding messages (e.g., postpartum cervical cancer screening). The results have important implications for efforts to improve postpartum care, considering historical underemphasis on the postnatal setting and the failure of existing systems to adequately deliver important aspects of care. PROMPTS was developed with this context in mind. It is not a standalone tool or substitute for formal healthcare; rather, it is intended to be a complement to the established healthcare infrastructure of Kenya, empowering patients as informed agents in their care and overcoming the limitations of often short and hurried clinical visits.

The impacts identified are also notable given the platform’s low cost. Considering the lifetime cost per enrollee estimated by Jacaranda Health, our results suggest that a mere 74 cents per patient can lead to a 17% relative increase (0.07/0.41; see **Table 5**) in the share of individuals receiving the guideline-recommended quantity of postnatal care and a 13% relative increase (0.07/0.53; see **Table 5**) in the share receiving postpartum family planning counseling.^36^

Further research is warranted in several areas. Given plans to scale up PROMPTS in settings outside of Kenya, it will be valuable to see how our findings generalize to contexts with different patterns of mobile phone penetration, health literacy, and MNH care seeking behavior. Future research would also be useful to identify subgroups that maximally benefit from the platform and to disentangle the extent to which the impacts we identify are driven by PROMPTS’ informational messages vs. appointment reminders vs. AI-enabled clinical helpdesk. Ongoing research is also examining the potential for synergies of patient-facing platforms like PROMPTS with health system level strategies to improve MNH outcomes.

## Data Availability

All data produced in the present study are available upon reasonable request to the authors for up to five years following the end of the study period, after which they will be destroyed.

## ACKNOWLEDGEMENTS

We are indebted to all the participants who took part in our study and generously shared their time with us. We are also grateful to our study enumerators for meticulously collecting the data on which our evaluation is based. Finally, we thank the full team at Jacaranda Health for their contributions to this study, particularly Jay Patel and Henry Njogu for carefully curating and sharing administrative data to guide our analyses; and Valerie Scott and Lujia Zhang (Harvard T.H. Chan School of Public Health) for their assistance in preparing the study data.

## REFERENCES

1 World Health Organization Sexual and Reproductive Health and Research Team. Trends in maternal mortality 2000 to 2020: estimates by WHO, UNICEF, UNFPA, World Bank Group and UNDESA/Population Division. 2023. https://www.who.int/publications/i/item/9789240068759

2 Roser M, Ritchie H, Dadonaite B. Child and Infant Mortality. Our World in Data. 2019. https://ourworldindata.org/child-mortality

3 United Nations Inter-Agency Group for Child Mortality Estimation. Levels & Trends in Child Mortality. 2023. https://data.unicef.org/wp-content/uploads/2023/01/UN-IGME-Child-Mortality-Report-2022_Final-online-version_9Jan.pdf

4 Kaye DK, Kakaire O, Osinde MO. Systematic review of the magnitude and case fatality ratio for severe maternal morbidity in sub-Saharan Africa between 1995 and 2010. BMC Pregnancy Childbirth. 2011 Sep 28;11:65. doi:10.1186/1471-2393-11-65

5 Haile TG, Gebregziabher D, Gebremeskel GG, Mebrahtom G, Aberhe W, Hailay A, et al. Prevalence of neonatal near miss in Africa: a systematic review and meta-analysis. International Health 2023; 0:1–10. doi.org/10.1093/inthealth/ihad034

6 Owolabi O, Riley T, Juma K, Mutua M, Pleasure ZH, Amo-Adjei J, et al. Incidence of maternal near-miss in Kenya in 2018: findings from a nationally representative cross-sectional study in 54 referral hospitals. Scientific Reports. 2020; 10:15181. doi.org/10.1038/s41598-020-72144-x

7 Nabwera HM, Wang D, Tongo OO, Andang’o PEA, Abdulkadir I, Ezeaka CV, et al. Burden of disease and risk factors for mortality amongst hospitalized newborns in Nigeria and Kenya. PLOS One. 2021 Jan 14;16(1):e0244109. doi:10.1371/journal.pone.0244109

8 The World Bank. Pregnant women receiving prenatal care (%) – Kenya. 2020. https://data.worldbank.org/indicator/SH.STA.ANVC.ZS?locations=KE

9 Atahigwa C, Kadengye DT, Iddi S, Abrams S, Van Rie A. Trends and determinants of health facility childbirth service utilization among mothers in urban slums of Nairobi, Kenya. Global Epidemiology. 2020; 2. doi.org/10.1016/j.gloepi.2020.100029

10 Kichamu G, Abisi JN, Karimurio L. Maternal and Child Health. Kenya Demographic and Health Survey. 2003. https://dhsprogram.com/pubs/pdf/fr151/09chapter09.pdf

11 Macharia PM, Joseph NK, Nalwadda GK, Mwilike B, Banke-Thomas A, Benova L, et al. Spatial variation and inequities in antenatal care coverage in Kenya, Uganda and mainland Tanzania using model-based geostatistics: a socioeconomic and geographical accessibility lens. BMC Pregnancy Childbirth. 2022; 22. doi.org/10.1186/s12884-022-05238-1

12 Kenya Department of Family Health. Healthy Mothers and Newborns; Guidelines for Postnatal Care. Kenya Ministry of Health. 2018. https://familyhealth.go.ke/wp-content/uploads/2018/02/Guidelines-for-postnatal-care-to-mothers-and-newborns-2016-18-12-2016B.pdf

13 Family Care International. Care-seeking during pregnancy, delivery and the postpartum period: a study in Homa Bay and Migori districts, Kenya. 2005.

14 Irving G, Neves AL, Dambha-Miller H, Oishi A, Tagashira H, Verho A, et al. International variations in primary care physician consultation time: a systematic review of 67 countries. BMJ Open. 2017 Nov 8;7(10):e017902. doi:10.1136/bmjopen-2017-017902.

15 Kruk ME, Gage AD, Arsenault C, Jordan K, Leslie HH, Roder-DeWan S, et al. High-quality health systems in the Sustainable Development Goals era: time for a revolution. Lancet Glob Health. 2018 Nov;6(11):e1196–e1252. doi:10.1016/S2214-109X(18)30386-3

16 Souza JP, Gülmezoglu AM, Vogel J, Carroli G, Lumbiganon P, Qureshi Z, et al. Moving beyond essential interventions for reduction of maternal mortality (the WHO Multicountry Survey on Maternal and Newborn Health): a cross-sectional study. The Lancet. 2013; 381(9879):1747–55. doi:10.1016/S0140-6736(13)60686-8

17 Kruk ME, Leslie HH, Verguet S, Mbaruku GM, Adanu RMK, Langer A. Quality of basic maternal care functions in health facilities of five African countries: an analysis of national health system surveys. The Lancet Global Health. 2016; 4(11):e845–e855. doi:10.1016/S2214-109X(16)30180-2

18 Sharma J, Leslie HH, Kundu F, Kruk ME. Poor Quality for Poor Women? Inequities in the Quality of Antenatal and Delivery Care in Kenya. PLOS One. 2017; 12(1):e0171236. doi:10.1371/journal.pone.0171236

19 Actis Danna V, Bedwell C, Wakasiaka S, Lavender T. Utility of the three-delays model and its potential for supporting a solution-based approach to accessing intrapartum care in low- and middle-income countries. A qualitative evidence synthesis. Global Health Action. 2020; 13(1):1819052. doi:10.1080/16549716.2020.1819052

20 Yunitasari E, Matos F, Zulkarnain H, Kumalasari DI, Kusumaningrum T, Putri TE, et al. Pregnant woman awareness of obstetric danger signs in developing country: systematic review. BMC Pregnancy and Childbirth. 2023; 23. 10.1186/s12884-023-05674-7

21 Phanice OM, Zachary MO. Knowledge of Obstetric Danger Signs among Pregnant Women Attending Antenatal Care Clinic at Health Facilities within Bureti Sub-County of Kericho County, Kenya. Research in Obstetrics and Gynecology. 2018; 6(1):16–21. doi:10.5923/j.rog.20180601

22 Carvalho OMC, Junior ABV, Augusto MCC, Leite ÁJM, Nobre RA, Bessa OAAC, et al. Delays in obstetric care increase the risk of neonatal near-miss morbidity events and death: a case-control study. BMC Pregnancy Childbirth. 2020; 20(1):437. doi:10.1186/s12884-020-03128-y

23 Tiruneh GA, Asaye MM, Solomon AA, Arega DT. Delays during emergency obstetric care and their determinants among mothers who gave birth in South Gondar zone hospitals, Ethiopia. A cross-sectional study design. Glob Health Action. 2021; 14(1):1953242. doi:10.1080/16549716.2021.1953242

24 Thaddeus S, Maine D. Too far to walk: maternal mortality in context. Social Science & Medicine. 1994; 38(8):1091–110. doi:10.1016/0277-9536(94)90226-7

25 Ersdal HL, Vossius C, Bayo E, Mduma E, Perlman J, Lippert A, et al. A one-day “Helping Babies Breathe” course improves simulated performance but not clinical management of neonates. Resuscitation. 2013; 84(10):1422–7. doi:10.1016/j.resuscitation.2013.04.005

26 Feyissa GT, Balabanova D, Woldie M. How Effective are Mentoring Programs for Improving Health Worker Competence and Institutional Performance in Africa? A Systematic Review of Quantitative Evidence. Journal of Multidisciplinary Healthcare. 2019; 12:989–1005. doi:10.2147/JMDH.S228951

27 Lassi ZS, Middleton PF, Bhutta ZA, Crowther C. Strategies for improving health care seeking for maternal and newborn illnesses in low- and middle-income countries: a systematic review and meta-analysis. Glob Health Action. 2016 May 10;9:31408. doi:10.3402/gha.v9.31408.

28 Mangham-Jefferies L, Pitt C, Cousens S, Mills A, Schellenberg J. Cost-effectiveness of strategies to improve the utilization and provision of maternal and newborn health care in low-income and lower-middle-income countries: a systematic review. BMC Pregnancy Childbirth. 2014;14(243). doi.org/10.1186/1471-2393-14-243.

29 Venkataramanan R, Subramanian SV, Alajlani M, Arvanitis TN. Effect of mobile health interventions in increasing utilization of Maternal and Child Health care services in developing countries: A scoping review. Digital Health. 2022; 8:1–12. doi:10.1177/20552076221143236

30 Feroz A, Perveen S, Aftab W. Role of mHealth applications for improving antenatal and postnatal care in low and middle income countries: a systematic review. BMC Health Services Research. 2017; 17(1):704. doi:10.1186/s12913-017-2664-7

31 Sondaal SFV, Browne JL, Amoakoh-Coleman M, Borgstein A, Miltenburg AS, Verwijs M, et al. Assessing the Effect of mHealth Interventions in Improving Maternal and Neonatal Care in Low- and Middle-Income Countries: A Systematic Review. PLOS ONE. 2016; 11(5):e0154664. doi.org/10.1371/journal.pone.0154664

32 Lee SH, Nurmatov UB, Nwaru BI, Mukherjee M, Grant L, Pagliari C. Effectiveness of mHealth interventions for maternal, newborn and child health in low- and middle-income countries: Systematic review and meta-analysis. Journal of Global Health. 2016; 6(1):010401. doi:10.7189/jogh.06.010401

33 Kabongo EM, Mukumbang FC, Delobelle P, Nicol E. Explaining the impact of mHealth on maternal and child health care in low- and middle-income countries: a realist synthesis. BMC Pregnancy and Childbirth. 2021; 21(1):196. doi:10.1186/s12884-021-03684-x

34 Alowais SA, Alghamdi SS, Alsuhebany N, Alqahtani T, Alshaya AI, Almohareb SN, et al. Revolutionizing healthcare: the role of artificial intelligence in clinical practice. BMC Med Educ. 2023 Sep 22;23(1):689. doi:10.1186/s12909-023-04698-z.

35 USAID. Artificial Intelligence in Global Health: Defining a Collective Path Forward. USAID Center for Innovation and Impact. 2022. https://www.usaid.gov/sites/default/files/2022-05/AI-in-Global-Health_webFinal_508.pdf

36 PROMPTS: PROmoting Mothers in Pregnancy and Postpartum Through SMS. Jacaranda Health. 2023. Available from: https://jacarandahealth.org/ypoagriw/2023/11/PROMPTS-BROCHURE-V4.pdf

37 Zhang W, Guo H, Ranganathan P, Patel J, Rajasekharan S, Danayak N, et al. A Continual Pre-training Approach to Tele-Triaging Pregnant Women in Kenya. Proceedings of the AAAI Conf. on Artificial Intelligence. 2023; 37(12):14620–14627. doi.org/10.1609/aaai.v37i12.26709

38 Anderson ML. Multiple Inference and Gender Differences in the Effects of Early Intervention: A Reevaluation of the Abecedarian, Perry Preschool, and Early Training Projects. 2008; 103(484):1481–1495.

39 Guidelines Review Committee. WHO recommendations on antenatal care for a positive pregnancy experience. World Health Organization. 2016. who.int/publications/i/item/9789241549912

40 Guidelines Review Committee. WHO recommendations on maternal and newborn care for a positive postnatal experience. World Health Organization. 2022. who.int/publications/i/item/9789240045989

41 Kenya National Bureau of Statistics. Kenya Demographic and Health Survey. 2014. https://www.dhsprogram.com/pubs/pdf/FR308/FR308.pdf

42 Datta SS, Ranganathan P, Sivakumar KS. A study to assess the feasibility of Text Messaging Service in delivering maternal and child healthcare messages in a rural area of Tamil Nadu, India. Australas Med J. 2014 Apr 30;7(4):175–80. doi:10.4066/AMJ.2014.1916

43 Gresh A, Cohen M, Anderson J, Glass N. Postpartum care content and delivery throughout the African continent: An integrative review. Midwifery. 2021 Jun;97:102976. doi:10.1016/j.midw.2021.102976

44 Guenther T, Nsona H, Makuluni R, Chisema M, Jenda G, Chimbalanga E, et al. Home visits by community health workers for pregnant mothers and newborns: coverage plateau in Malawi. J Glob Health. 2019 Jun;9(1):010808. doi:10.7189/jogh.09.010808.

45 Subramanian L, Murthy S, Bogam P, Yan SD, Marx Delaney M, Goodwin CDG, et al. Just-in-time postnatal education programmes to improve newborn care practices: needs and opportunities in low-resource settings. BMJ Glob Health. 2020 Jul;5(7):e002660. doi:10.1136/bmjgh-2020-002660.

46 Lau YK, Cassidy T, Hacking D, Brittain K, Haricharan HJ, Heap M. Antenatal health promotion via short message service at a Midwife Obstetrics Unit in South Africa: a mixed methods study. BMC Pregnancy Childbirth. 2014 Aug 21;14:284. doi:10.1186/1471-2393-14-284.

47 Soubeiga D, Gauvin L, Hatem MA, Johri M. Birth Preparedness and Complication Readiness (BPCR) interventions to reduce maternal and neonatal mortality in developing countries: systematic review and meta-analysis. BMC Pregnancy and Childbirth. 2014; 14:129. doi:10.1186/1471-2393-14-129

48 Fedha, T. Impact of Mobile Telephone on Maternal Health Service Care: A Case of Njoro Division. Open Journal of Preventive Medicine. 2014; 4:365–376.

49 Lund S, Nielsen BB, Hemed M, Boas IM, Said A, Said K, et al. Mobile phones improve antenatal care attendance in Zanzibar: a cluster randomized controlled trial. BMC Pregnancy Childbirth. 2014 Jan 17;14:29. doi:10.1186/1471-2393-14-29.

50 Adanikin AI, Awoleke JO, Adeyiolu A. Role of reminder by text message in enhancing postnatal clinic attendance. Int J Gynaecol Obstet. 2014 Aug;126(2):179–80. doi:10.1016/j.ijgo.2014.02.009.

51 Soucat A, Levy-Bruhl D, De B, Gbedonou P, Lamarque JP, Bangoura O, et al. Affordability, cost-effectiveness and efficiency of primary health care: the Bamako Initiative experience in Benin and Guinea. Int J Health Plann Manag 1997;12(Suppl 1):S81–S108.

52 Hutchinson P, Lance P, Guilkey DK, Shahjahan M, Haque S. Measuring the cost-effectiveness of a national health communication program in rural Bangladesh. J Health Commun. 2006;11(Suppl 2):91–121.

53 Pape UJ, Delius A, Khandelwal R, Gupta R. Socioeconomic Impacts of COVID-19 in Kenya. World Bank. 2021. http://hdl.handle.net/10986/35961

