## Supplementary Materials for "Empowering patients across the maternal-newborn care continuum: A cluster randomized controlled trial testing a digital health platform in Kenya"

**Appendix S1:** Technical overview of facility level randomization.

Using the randomize command in Stata, we iterated through 1:1 treatment-to-control randomization assignments until the corresponding balance p-value exceeded 0.7, with a maximum of 2,000 iterations permitted. The balance p-value accounted for facility-level caesarean birth frequency (i.e., proportion of total deliveries conducted by caesarean section) and perinatal mortality (i.e., proportion of total stillbirths and live births ending in stillbirth or neonatal death within one week).

**Appendix S2:** Detailed description of knowledge, preparedness, and danger sign care seeking outcome measures.

*Knowledge Domain*

- Knowledge of labor signs was compared against a list of 7 actual labor signs, including changes in discharge, contractions, heavy bleeding, lower abdominal pain, lower back pain, urgency to go to the toilet, and water breaking.
- Danger sign knowledge questions inquired whether a respondent would seek immediate medical care or watchfully wait in response to several potentially severe symptoms. Specifically, 6 antenatal symptoms (back pain, blurred vision, decreased fetal movement, heartburn, nausea/vomiting, and vaginal bleeding), 3 postpartum symptoms (blurred vision, chest pain, and vaginal bleeding), and 3 neonatal symptoms (fever, yellow eyes, and yellow/thin stool) were assessed.

*Preparedness Domain*

- The total number of items reported to have been completed in preparation for childbirth was compared against a list of eight items, including asking family or friends to help with childcare, buying baby clothes, choosing a hospital, discussing a birth plan, packing a bag, planning transportation, purchasing insurance, and saving money.

*Danger Sign Care Seeking Domain*

- Participants were asked whether they experienced any of:
    - 10 antenatal danger signs: blurred vision, breathing difficulty, contractions before 37 weeks, convulsions/loss of consciousness, decreased/absent fetal movements, fever, leaking of fluid before 37 weeks, severe abdominal pain, severe headache, and vaginal bleeding/discharge with foul odor.
    - 9 postpartum danger signs: blurred vision, breathing difficulty, calf pain/redness/swelling, chest pain, convulsions/loss of consciousness, fever, heavy/suddenly increased vaginal bleeding, severe abdominal pain, and severe headache.
    - 9 neonatal danger signs: convulsions/fits, fast or difficult breathing, fever, hypothermia, inability to feed/poor feeding, regurgitating with each feeding, umbilical redness/drainage, weakness/lethargy, and yellow eyes/soles of extremities.

**SUPPLEMENTAL TABLES**

**Table S1:** Attrition in Baseline Study Sample During Antenatal and Postpartum Follow-Up

| Attrition Variable | Control | Treated | *p*-value of Difference [a] |
|---|---|---|---|
| Attrition from Baseline to Antenatal Follow-Up [b] | 123 / 1813 (6.8%) | 156 / 1865 (8.4%) | 0.19 |
| Attrition from Baseline to Postpartum Follow-Up [c] | 321 / 2997 (10.7%) | 298 / 3131 (9.5%) | 0.24 |

[a] p-values from individual-level regressions adjusted for a treatment indicator and recruitment-facility-level normal vaginal birth volume tertile at baseline; cluster robust standard errors clustered by recruitment facility.
[b] Attrition among targeted and eligible participants at antenatal follow-up.
[c] Attrition among targeted and eligible participants at postpartum follow-up.

**Table S2:** Baseline Characteristics Among Eligible and Consented Cohort of Pregnant Women Recruited During ANC Visits to Study Facilities Who Completed Antenatal Follow-Up

| Baseline Characteristic | Control (N = 1,690) | Treated (N = 1,709) |
|---|---|---|
| Age (years) | 26.05 (5.87) | 26.24 (5.77) |
| Secondary school education or higher | 65.9% | 68.5% |
| Read Kiswahili or English without difficulty | 94.4% | 96.1% |
| Married or living together | 82.6% | 83.7% |
| Size of household | 3.93 (2.03) | 3.58 (1.83) |
| Own land | 47.9% | 38.2% |
| Improved source of drinking water | 68.7% | 71.8% |
| Improved sanitation facility | 97.6% | 98.2% |
| Mode of travel to hospital: motor vehicle | 69.8% | 68.1% |
| Time to travel from home to health facility (minutes) | 24.30 (18.80) | 23.10 (18.49) |
| Work for pay in last week | 24.6% | 25.4% |
| Easy access to Ksh 2000 if treatment for illness needed in household | 27.8% | 28.9% |
| Own mobile phone | 90.0% | 93.3% |
| Use mobile phone to send text messages frequently/daily | 34.2% | 40.1% |
| Previous receipt of text message offering pregnancy advice from county | 3.5% | 4.9% |
| Gestational age (weeks) | 27.18 (5.19) | 27.09 (5.12) |
| Any prior prenatal care visit | 75.1% | 74.3% |
| # prenatal care visits for current pregnancy | 1.54 (1.32) | 1.49 (1.29) |
| Fraction of knowledge questions answered correctly | 0.67 (0.18) | 0.69 (0.18) |
| High-risk pregnancy | 22.1% | 17.5% |
| # of total pregnancies, including current pregnancy | 2.42 (1.46) | 2.28 (1.41) |
| Previous high-risk pregnancy | 39.9% | 44.5% |
| PHQ-2 score | 1.56 (1.60) | 1.61 (1.60) |

Abbreviations: ANC, antenatal care; PHQ-2, Patient Health Questionnaire-2

Continuous variables summarized by sample mean and standard deviation: mean (SD); binary variables summarized by sample mean as a %.

**Table S3:** Baseline Characteristics Among Eligible and Consented Cohort of Pregnant Women Recruited During ANC Visits to Study Facilities Who Completed Postpartum Follow-Up

| Baseline Characteristic | Control (N = 2,676) | Treated (N = 2,833) |
|---|---|---|
| Age (years) | 26.08 (5.91) | 26.37 (5.80) |
| Secondary school education or higher | 64.6% | 67.6% |
| Read Kiswahili or English without difficulty | 95.1% | 96.5% |
| Married or living together | 81.9% | 82.9% |
| Size of household | 3.99 (2.05) | 3.61 (1.84) |
| Own land | 47.8% | 37.4% |
| Improved source of drinking water | 68.8% | 70.8% |
| Improved sanitation facility | 97.1% | 98.3% |
| Mode of travel to hospital: motor vehicle | 70.1% | 69.3% |
| Time to travel from home to health facility (minutes) | 24.33 (18.26) | 22.94 (17.57) |
| Work for pay in last week | 22.3% | 22.9% |
| Easy access to Ksh 2000 if treatment for illness needed in household | 27.6% | 28.2% |
| Own mobile phone | 89.3% | 92.1% |
| Use mobile phone to send text messages frequently/daily | 33.6% | 40.7% |
| Previous receipt of text message offering pregnancy advice from county | 4.2% | 6.3% |
| Gestational age (weeks) | 29.75 (6.17) | 29.83 (6.14) |
| Any prior prenatal care visit | 81.7% | 81.4% |
| # prenatal care visits for current pregnancy | 1.93 (1.47) | 2.00 (1.56) |
| Fraction of knowledge questions answered correctly | 0.67 (0.18) | 0.69 (0.18) |
| High-risk pregnancy | 23.7% | 19.7% |
| # of total pregnancies, including current pregnancy | 2.43 (1.48) | 2.30 (1.39) |
| Previous high-risk pregnancy | 40.5% | 44.0% |
| PHQ-2 score | 1.50 (1.57) | 1.62 (1.60) |

Abbreviations: ANC, antenatal care; PHQ-2, Patient Health Questionnaire-2

Continuous variables summarized by sample mean and standard deviation: mean (SD); binary variables summarized by sample mean as a %

**Table S4:** Completed Consolidated Standards of Reporting Trials (CONSORT) checklist of information to include when reporting a randomized trial, with extensions for cluster-randomized trials

| Section/Topic | Item No | Standard Checklist item | Extension for Cluster Designs | Reported in… |
|---|---|---|---|---|
| **Title and abstract** | | | | |
| | 1a | Identification as a randomized trial in the title | Identification as a cluster randomized trial in the title | Title Page |
| | 1b | Structured summary of trial design, methods, results, and conclusions (for specific guidance see CONSORT for abstracts) | See Table S5 | Abstract |
| **Introduction** | | | | |
| Background and objectives | 2a | Scientific background and explanation of rationale | Rationale for using a cluster design | Introduction (para. 1-7) |
| | 2b | Specific objectives or hypotheses | Whether objectives pertain to the cluster level, the individual participant level, or both | Introduction (para. 7) |
| **Methods** | | | | |
| Trial design | 3a | Description of trial design (such as parallel, factorial) including allocation ratio | Definition of cluster and description of how the design features apply to the clusters | Methods (para. 1) |
| | 3b | Important changes to methods after trial commencement (such as eligibility criteria), with reasons | | N/A |
| Participants | 4a | Eligibility criteria for participants | Eligibility criteria for clusters | Methods > Procedures > Health Facility Eligibility and Methods > Procedures > Recruitment and Eligibility |
| | 4b | Settings and locations where the data were collected | | Methods > Procedures > Recruitment and |

| | | | | |
|---|---|---|---|---|
| | | | | Eligibility; Methods > Procedures Data Collection; Methods > Procedures > Enrollment in PROMPTS; Figure 1 |
| Interventions | 5 | The interventions for each group with sufficient details to allow replication, including how and when they were actually administered | Whether interventions pertain to the cluster level, the individual participant level, or both | Methods (para. 2) and Methods > Intervention |
| Outcomes | 6a | Completely defined pre-specified primary and secondary outcome measures, including how and when they were assessed | Whether outcome measures pertain to the cluster level, the individual participant level, or both | Methods > Outcomes |
| | 6b | Any changes to trial outcomes after the trial commenced, with reasons | | Methods > Outcomes |
| Sample size | 7a | How sample size was determined | Method of calculation, number of clusters(s) (and whether equal or unequal cluster sizes are assumed), cluster size, a coefficient of intracluster correlation (ICC or k), and an indication of its uncertainty | Methods > Statistical Analysis > Sample Size and Power |
| | 7b | When applicable, explanation of any interim analyses and stopping guidelines | | N/A |
| Randomization | | | | |
| Sequence generation | 8a | Method used to generate the random allocation sequence | | Methods > Randomization and Appendix S1 |
| | 8b | Type of randomization; details of any restriction (such as blocking and block size) | Details of stratification or matching if used | Methods > Randomization and Appendix S1 |

| | | | | |
|---|---|---|---|---|
| Allocation concealment mechanism | 9 | Mechanism used to implement the random allocation sequence (such as sequentially numbered containers), describing any steps taken to conceal the sequence until interventions were assigned | Specification that allocation was based on clusters rather than individuals and whether allocation concealment (if any) was at the cluster level, the individual participant level, or both | Methods > Randomization |
| Implementation | 10 | Who generated the random allocation sequence, who enrolled participants, and who assigned participants to interventions | Replaced by 10a, 10b, and 10c | |
| | 10a | | Who generated the random allocation sequence, who enrolled clusters, and who assigned clusters to interventions | Methods > Randomization; Methods > Procedures > Health Facility Eligibility; Appendix S1 |
| | 10b | | Mechanism by which individual participants were included in clusters for the purposes of the trial (such as complete enumeration, random sampling) | Methods (para. 2) and Methods > Procedures > Recruitment and Eligibility |
| | 10c | | From whom consent was sought (representatives of the cluster, or individual cluster members, or both) and whether consent was sought before or after randomization | Methods > Procedures > Data Collection; Methods > Procedures > Enrollment in PROMPTS; Methods > Ethics and Safety Statement |
| Blinding | 11a | If done, who was blinded after assignment to interventions (for example, participants, care providers, those assessing outcomes) and how | | N/A |

| | | | | |
|---|---|---|---|---|
| | 11b | If relevant, description of the similarity of interventions | | N/A |
| Statistical methods | 12a | Statistical methods used to compare groups for primary and secondary outcomes | How clustering was taken into account | Methods > Statistical Analysis |
| | 12b | Methods for additional analyses, such as subgroup analyses and adjusted analyses | | Methods > Statistical Analysis |
| **Results** | | | | |
| Participant flow (a diagram is strongly recommended) | 13a | For each group, the numbers of participants who were randomly assigned, received intended treatment, and were analyzed for the primary outcome | For each group, the numbers of clusters that were randomly assigned, received intended treatment, and were analyzed for the primary outcome | Results > Sample Attrition and Characteristics (para. 1) and Figure 2 |
| | 13b | For each group, losses and exclusions after randomization, together with reasons | For each group, losses and exclusions for both clusters and individual cluster members | Results > Sample Attrition and Characteristics (para. 1); Results > Intervention Fidelity; Tables 3 and S1 |
| Recruitment | 14a | Dates defining the periods of recruitment and follow-up | | Methods > Procedures > Data Collection and Figure 2 |
| | 14b | Why the trial ended or was stopped | | Methods > Procedures > Data Collection |
| Baseline data | 15 | A table showing baseline demographic and clinical characteristics for each group | Baseline characteristics for the individual and cluster levels as applicable for each group | Results > Sample Attrition and Characteristics (para. 2) and Tables 2, S2-S3 |
| Numbers analyzed | 16 | For each group, number of participants (denominator) included in each analysis and whether the | For each group, number of clusters included in each analysis | Methods > Statistical Analysis; Results > Sample Attrition and |

| | | | | |
|---|---|---|---|---|
| | | analysis was by original assigned groups | | Characteristics (para. 1); and Tables S2-S3 |
| Outcomes and estimation | 17a | For each primary and secondary outcome, results for each group, and the estimated effect size and its precision (such as 95% confidence interval) | Results at the individual or cluster level as applicable and a coefficient of intracluster correlation (ICC or k) for each primary outcome | Results > Intervention Impact; Tables 4 and 5; Figures 4 and 5; ICC not reported |
| | 17b | For binary outcomes, presentation of both absolute and relative effect sizes is recommended | | Results > Intervention Impact and Discussion > Implications and Future Directions |
| Ancillary analyses | 18 | Results of any other analyses performed, including subgroup analyses and adjusted analyses, distinguishing pre-specified from exploratory | | Results > Intervention Impact; Tables 4 and 5; Figure 5 |
| Harms | 19 | All important harms or unintended effects in each group (for specific guidance see CONSORT for harms) | | N/A |
| **Discussion** | | | | |
| Limitations | 20 | Trial limitations, addressing sources of potential bias, imprecision, and, if relevant, multiplicity of analyses | | Discussion > Strengths and Limitations |
| Generalizability | 21 | Generalizability (external validity, applicability) of the trial findings | Generalizability to clusters and/or individual participants (as relevant) | Discussion > Implications and Future Directions |
| Interpretation | 22 | Interpretation consistent with results, balancing benefits and harms, and considering other relevant evidence | | Discussion (para. 1-8) |
| **Other information** | | | | |
| Registration | 23 | Registration number and name of trial registry | | Abstract > Trial Registration and Methods (para. 3) |

| | | | | |
|---|---|---|---|---|
| Protocol | 24 | Where the full trial protocol can be accessed, if available | | Methods > Procedures; additional information available upon request to authors |
| Funding | 25 | Sources of funding and other support (such as supply of drugs), role of funders | | Disclosed to Journal |

Citation: Schulz KF, Altman DG, Moher D, for the CONSORT Group. CONSORT 2010 Statement: updated guidelines for reporting parallel group randomised trials. BMC Medicine. 2010;8:18.

**Table S5:** Completed CONSORT checklist for abstracts to reports of cluster randomized trials, with extensions for cluster-randomized trials

| Section/Topic | Standard Checklist Item | Extension for Cluster Designs | Reported in… |
|---|---|---|---|
| Title | Identification of the study as randomized | Identification of study as cluster randomized | Title page |
| Trial design | Description of the trial design (for example, parallel, cluster, non-inferiority) | | Abstract > Methods and Findings |
| Methods: | | | |
| Participants | Eligibility criteria for participants and the settings where the data were collected | Eligibility criteria for clusters | Insufficient room in abstract; reported in main text in Methods > Procedures > Health Facility Eligibility and Methods > Procedures > Recruitment and Eligibility |
| Interventions | Interventions intended for each group | | Abstract > Methods and Findings |
| Objective | Specific objective or hypothesis | Whether objective or hypothesis pertains to the cluster level, the individual participant level, or both | Abstract > Methods and Findings |
| Outcome | Clearly defined primary outcome for this report | Whether the primary outcome pertains to the cluster level, the individual participant level or both | Abstract > Methods and Findings |
| Randomization | How participants were allocated to interventions | How clusters were allocated to interventions | Insufficient room in abstract; reported in main text in Methods > Procedures > Randomization |

| | | | |
|---|---|---|---|
| Blinding (masking) | Whether or not participants, care givers, and those assessing the outcomes were blinded to group assignment | | Abstract > Methods and Findings |
| Results: | | | |
| Numbers randomized | Number of participants randomized to each group | Number of clusters randomized to each group | Number of clusters randomized to each group implied in Abstract > Methods and Findings |
| Numbers analyzed | Number of participants analyzed in each group | Number of clusters analyzed in each group | Insufficient room in abstract to disaggregate by group; reported in main text in Results > Sample Attrition and Characteristics |
| Outcome | For the primary outcome, a result for each group and the estimated effect size and its precision | Results at the cluster or individual level as applicable for each primary outcome | Abstract > Methods and Findings |
| Harms | Important adverse events or side effects | | N/A |
| Conclusions | General interpretation of the results | | Abstract > Conclusions |
| Trial registration | Registration number and name of trial register | | Abstract > Trial Registration |
| Funding | Source of funding | | Disclosed to Journal |

Citation: Campbell MK, Piaggio G, Elbourne DR, Altman DG; CONSORT Group. Consort 2010 statement: extension to cluster randomised trials. BMJ. 2012 Sep 4;345:e5661.
